# Predictors of SARS-CoV-2 infection in a multi-ethnic cohort of United Kingdom healthcare workers: a prospective nationwide cohort study (UK-REACH)

**DOI:** 10.1101/2021.12.16.21267934

**Authors:** Christopher A. Martin, Daniel Pan, Carl Melbourne, Lucy Teece, Avinash Aujayeb, Rebecca F. Baggaley, Luke Bryant, Sue Carr, Bindu Gregary, Amit Gupta, Anna L. Guyatt, Catherine John, I Chris McManus, Joshua Nazareth, Laura B. Nellums, Rubina Reza, Sandra Simpson, Martin D. Tobin, Katherine Woolf, Stephen Zingwe, Kamlesh Khunti, Keith R. Abrams, Laura J. Gray, Manish Pareek

**Affiliations:** Department of Respiratory Sciences, University of Leicester, Leicester, UK; Department of Infection and HIV Medicine, University Hospitals of Leicester NHS Trust, Leicester, UK; Genetic Epidemiology Research Group, Department of Health Sciences, University of Leicester, Leicester, UK; Biostatistics Research Group, Department of Health Sciences, University of Leicester, Leicester, UK; Respiratory Department, Northumbria Specialist Emergency Care Hospital; Department of Nephrology, University Hospitals of Leicester NHS Trust, Leicester, UK; General Medical Council, London, UK; Lancashire Clinical Research Facility, Royal Preston Hospital; Oxford University Hospitals NHS Foundation Trust, Oxford, UK; Department of Health Sciences, University of Leicester, Leicester, UK; University College London Medical School, London, UK; Population and Lifespan Sciences, School of Medicine, University of Nottingham, Nottingham, UK; Centre for Research & Development, Derbyshire Healthcare NHS Foundation Trust, Derby, UK; Nottinghamshire Healthcare NHS Foundation Trust, Nottingham, UK; Research and Development Department, Berkshire Healthcare NHS Foundation Trust, Bracknell, UK; Diabetes Research Centre, University of Leicester, Leicester, UK; Department of Statistics, University of Warwick, UK; Chief investigator; University of Leicester; University of Nottingham; University College London; Oxford University Hospitals; University of York; University Hospital Leicester; University of Edinburgh; University of Swansea

## Abstract

**Introduction:** Healthcare workers (HCWs), particularly those from ethnic minority groups, have been shown to be at disproportionately higher risk of infection with severe acute respiratory syndrome coronavirus-2 (SARS-CoV-2) compared to the general population. However, there is insufficient evidence on how demographic and occupational factors influence infection risk among ethnic minority HCWs.

**Methods:** We conducted a cross-sectional analysis using data from the United Kingdom Research study into Ethnicity And COVID-19 Outcomes in Healthcare workers (UK-REACH) cohort study. We used logistic regression to examine associations of demographic, household and occupational predictor variables with SARS-CoV-2 infection (defined by PCR, serology or suspected COVID-19) in a diverse group of HCWs.

**Results:** 2,496 of the 10,772 HCWs (23.2%) who worked during the first UK national lockdown in March 2020 reported previous SARS-CoV-2 infection. In an adjusted model, demographic and household factors associated with increased odds of infection included younger age, living with other key workers and higher religiosity. Important occupational risk factors associated with increased odds of infection included attending to a higher number of COVID-19 positive patients (aOR 2.49, 95%CI 2.03–3.05 for ≥21 patients per week vs none), working in a nursing or midwifery role (1.35, 1.15– 1.58, compared to doctors), reporting a lack of access to personal protective equipment (1.27, 1.15 – 1.41) and working in an ambulance (1.95, 1.52–2.50) or hospital inpatient setting (1.54, 1.37 – 1.74). Those who worked in Intensive Care Units were less likely to have been infected (0.76, 0.63–0.90) than those who did not. Black HCWs were more likely to have been infected than their White colleagues, an effect which attenuated after adjustment for other known predictors.

**Conclusions:** We identified key sociodemographic and occupational risk factors associated with SARS-CoV-2 infection amongst UK HCWs, and have determined factors that might contribute to a disproportionate odds of infection in HCWs from Black ethnic groups. These findings demonstrate the importance of social and occupational factors in driving ethnic disparities in COVID-19 outcomes, and should inform policies, including targeted vaccination strategies and risk assessments aimed at protecting HCWs in future waves of the COVID-19 pandemic.

**Trial registration:** ISRCTN 11811602

## Introduction

The first patients with coronavirus disease 2019 (COVID-19) in the United Kingdom (UK) were identified in late January 2020.^1^ Thousands of healthcare workers (HCWs) in the UK have since been infected with Severe Acute Respiratory Syndrome Coronavirus-2 (SARS-CoV-2). A report by Public Health England suggested that early in the pandemic, up to 73% of infections in HCWs were due to nosocomial transmission.^2^ However, there remains insufficient evidence around key predictors of infection in HCWs, and particularly what is driving reported ethnic disparities in infection risk. A recent study in the USA of over 24,000 HCWs found community exposures to be important in driving SARS-CoV-2 seropositivity but found no occupational predictors of infection,^3^ whereas a study of the workforce in one hospital in the UK found occupational factors to be important predictors of SARS-CoV-2 seropositivity.^4^ Specific risk factors contributing to an increased risk of COVID-19 among HCWs from some ethnic minority groups are also poorly understood.^5^

We sought to address these knowledge gaps using data from the national United Kingdom Research study into Ethnicity and COVID-19 diagnosis and outcomes in Healthcare workers (UK-REACH) longitudinal cohort study, which is amongst the largest UK HCW cohort studies and is unique in the richness of its dataset and the ethnic diversity of its participants. Specifically, we sought to determine predictors of infection in UK HCWs and whether any disproportionate risks of infection in HCWs from ethnic minority groups might be explained by such predictors.

## Methods

### Overview

UK-REACH is a programme of work aiming to determine the impact of the COVID-19 pandemic on UK HCWs, and establish whether, and to what degree, this differs according to ethnicity. This cross sectional analysis uses data from the baseline questionnaire of the prospective nationwide cohort study, administered between December 2020 and March 2021.

Details of the study design, sampling and measures included in the baseline questionnaire can be found in the study protocol^6^ and the data dictionary (https://www.uk-reach.org/data-dictionary).

### Study population

We recruited individuals aged 16 years or over, living in the UK and employed as HCWs or ancillary workers in a healthcare setting and/or registered with one of the following UK professional regulatory bodies: the General Medical Council, Nursing and Midwifery Council, General Dental Council, Health and Care Professions Council, General Optical Council, General Pharmaceutical Council, or the Pharmaceutical Society of Northern Ireland.

### Recruitment

We asked professional regulators to distribute emails to their registrants embedded with a hyperlink to the study website. The sample was supplemented by direct recruitment of participants through participating healthcare trusts, and advertising on social media and in newsletters. Those interested could create a user profile, read the participant information sheet and, if they were willing, sign an online consent form. After providing consent, participants were asked to complete the questionnaire.

Participation rates at each stage are reported as recommended by the Checklist for Reporting Results of Internet E-Surveys (CHERRIES).^7,8^

### Outcome measures

Our primary outcome was SARS-CoV-2 infection, as determined by the self-reporting of either a positive polymerase chain reaction (PCR) assay for SARS-CoV-2 or a positive anti-SARS-CoV-2 serology assay. In addition, to ensure those that who acquired infection prior to widespread testing availability were not excluded, in those who had never been tested by PCR or serology, we included those individuals whose infection status was based on whether they, or another healthcare professional, suspected them of having had COVID-19 (see Supplementary Table 1 for details).

### Predictor variables

Our primary exposure of interest was self-reported ethnicity, categorised using the UK’s Office for National Statistics (ONS) 5- and 18-level ethnic group categories.^9^ For the main analysis, ethnicity was categorised into five broad ethnic groups (White, Asian, Black, Mixed and Other) to maximise the statistical power to test differences between groups. To ensure that we did not overlook important findings through collapsing ethnicity into broad groups, we also conducted additional analyses using 18 ethnicity categories.

Other variables potentially associated with the outcome were selected *a priori* based on the existing literature and expert opinion. These comprised:

- Demographic characteristics (age, sex).
- Occupational factors (job role, area of work, number of confirmed/suspected COVID-19 patients seen per week with physical contact, sharing transport to work with those outside of the household, access to personal protective equipment [PPE], exposure to aerosol generating procedures [AGPs], hours worked per week and night shift frequency).
- Household/residential/social factors (index of multiple deprivation [IMD, the official measure of relative deprivation for small areas of England, expressed as quintiles],^10^ number of occupants in household, types of social contact [remote only, face-to-face with social distancing or with physical contact], whether participants were cohabiting with another key worker (defined as someone expected to work during lockdown restrictions), and whether a participant’s accommodation contained spaces that were shared with other households.
- Comorbidities (diabetes and immunosuppression) that might be associated with acquiring infection (as opposed to risk of severe disease).
- Smoking status.
- UK region of workplace.
- Religiosity (i.e., how important a participant felt religion was in their daily life) and migration status were included to examine whether these might mediate any differences in infection risk found between ethnic groups. We included religiosity rather than religion as it was felt that the relative importance of religion, and thus the inclination to attend religious gatherings/places of worship during the pandemic, was more important in terms of acquisition of SARS-CoV-2 infection than the specific religion of a participant.

A description of each variable and how it was derived from questionnaire responses can be found in Supplementary Table 2.

### Statistical analysis

We excluded those with missing data for the primary exposure (ethnicity) and outcome of interest (SARS-CoV-2 infection) from all analyses. Occupational variables used in the analysis reflect the participants’ occupational circumstances during the weeks after implementation of the first national lockdown in the UK (which began on 23^rd^ March 2020). Therefore, in the main analysis we excluded those not working during this time. We undertook an additional analysis examining demographic and home factors only (leaving workplace region in the model as a proxy for region in which the participant lived) in all participants.

We summarised categorical variables as frequency and percentage, and non-normally distributed continuous variables as median (interquartile range [IQR]). We compared demographic, household and occupational factors between ethnic groups using chi-square tests for categorical data and Kruskal-Wallis tests for continuous data.

We used univariable and multivariable logistic regression to determine unadjusted and adjusted associations of the variables described above with self-reported history of SARS-CoV-2 infection and report results as unadjusted and adjusted odds ratios (ORs and aORs) and 95% confidence intervals (95%CIs).

Multiple imputation was used to impute missing data in these logistic regression models. The imputation models included all variables used in the final analyses bar those being imputed. Rubin’s Rules were used to combine the parameter estimates and standard errors from 10 imputations into a single set of results.^11^ Although indices of deprivation are available for UK countries outside England, it is recognised that these are not directly comparable with English IMD.^12^ We therefore elected to code IMD as missing for those outside England and impute the missing information.

We undertook three sensitivity analyses. Firstly, an analysis was conducted including only HCWs who had been tested for evidence of SARS-CoV-2 infection by PCR or serology. Secondly, to account for the fact that antibodies to SARS-CoV-2 may be induced by vaccination, we recoded those determined to have been infected solely by a positive serology assay as uninfected if their antibody result date was both valid (as determined by its temporal association with questionnaire completion date) and later than their vaccination date. Thirdly, and finally, we undertook a complete case analysis.

To investigate the extent to which differences in infection risk by ethnic group could be explained by other related predictors, we generated a base logistic regression model additionally adjusted for age and sex, and sequentially adjusted first for household/social/residential factors, second adding occupational risk factors, third adding health factors, fourth adding work region, and finally adding religiosity and migration status.

All analyses were conducted using Stata 17 (StataCorp. 2021. Stata Statistical Software: Release 17. College Station, TX: StataCorp LLC.)

### Ethical approval

The study was approved by the Health Research Authority (Brighton and Sussex Research Ethics Committee; ethics reference: 20/HRA/4718). All participants gave informed consent.

### Involvement and engagement

We worked closely with a Professional Expert Panel of HCWs from a range of ethnic backgrounds, healthcare occupations, and sexes, as well as with national and local organisations (see study protocol).^6^

### Role of the funding source

The funders had no role in study design, data collection, data analysis, interpretation, or writing of the report.

## Results

### Cohort recruitment and formation of the analysis sample

The recruitment of the cohort has been described previously and details, including response rates, are shown in Figure 1.^13,14^ Briefly, 15,119 HCWs started the questionnaire, of whom 1,858 were excluded from the current analysis as they did not provide their ethnicity, and 720 were excluded due to a lack of outcome data. Therefore, 12,541 HCWs formed the analysis sample, 1,769 of whom were not working during lockdown and therefore were not included in analyses of occupational determinants of infection.

**Figure 1.**
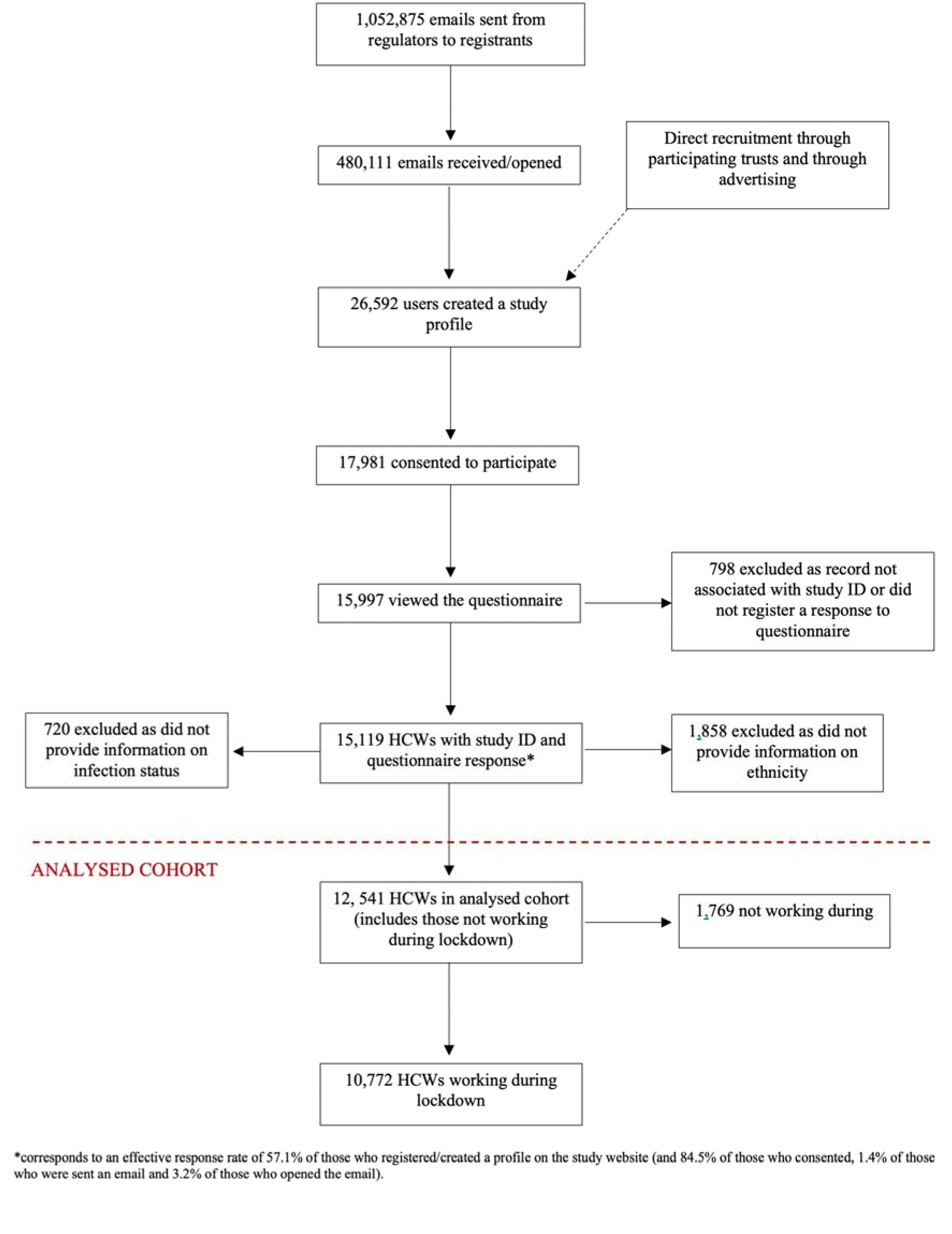
Formation of the analysed cohort.

### Description of the analysed cohort

A description of the cohort is shown in Table 1. With reference to the cohort who were working during lockdown (n=10,772), the majority were female (75.1%) with a median age of 45 (IQR 35 – 54). Approximately 30% were from ethnic minority groups (19.1% Asian, 4.3% Black, 4.1% Mixed, 2.1% Other).

**Table 1.** Description of the two analysed cohorts.

| Variable | Excluding those not working during lockdown | All Participants with non-missing ethnicity and infection status |
| --- | --- | --- |
|  | Total<br>N=10,772 | Total<br>N=12,541 |
| **Demographic and household factors** |  |  |
| **Age**, med(IQR) | 45 (35 – 54) | 45 (34 – 54) |
| Missing | 54 (0.5%) | 68 (0.5%) |
| **Sex** |  |  |
| Male | 2660 (24.7%) | 2977 (23.7%) |
| Female | 8089 (75.1%) | 9535 (76.0%) |
| Missing | 23 (0.2%) | 29 (0.2%) |
| **Ethnicity** |  |  |
| White | 7583 (70.4%) | 8795 (70.1%) |
| Asian | 2057 (19.1%) | 2418 (19.3%) |
| Black | 462 (4.3%) | 535 (4.3%) |
| Mixed | 446 (4.1%) | 529 (4.2%) |
| Other | 224 (2.1%) | 264 (2.1%) |
| Missing | 0 (0.0%) | 0 (0.0%) |
| **Migration status** |  |  |
| Born in UK | 7901 (73.5%) | 9171 (73.1%) |
| Born abroad | 2847 (26.5%) | 3341 (26.6%) |
| Missing | 24 (0.2%) | 29 (0.2%) |
| **Religiosity** |  |  |
| Not religious or not important | 6085 (56.5%) | 7043 (56.2%) |
| Fairly important | 2268 (21.1%) | 2637 (21.0%) |
| Very important | 1064 (9.9%) | 1238 (9.9%) |
| Extremely important | 1124 (10.4%) | 1335 (10.7%) |
| Missing | 231 (2.1%) | 288 (2.3%) |
| **Household size**, med (IQR) | 2 (1 – 3) | 2 (1 – 3) |
| Missing | 8 (0.1%) | 11 (0.1%) |
| **Cohabitation** |  |  |
| Does not live with other key workers | 5571 (51.7%) | 6574 (52.4%) |
| Lives with other key workers | 5145 (47.8%) | 5892 (47.0%) |
| Missing | 56 (0.5%) | 75 (0.6%) |
| **Accommodation** |  |  |
| Does not have shared spaces | 8807 (81.8%) | 10,248 (81.7%) |
| Has shared spaces | 1905 (17.7%) | 2221 (17.7%) |
| Missing | 60 (0.6%) | 72 (0.6%) |
| **Index of multiple deprivation quintile** |  |  |
| 1 (most deprived) | 956 (8.9%) | 1112 (8.9%) |
| 2 | 1597 (14.8%) | 1840 (14.7%) |
| 3 | 1944 (18.1%) | 2301 (18.4%) |
| 4 | 2312 (21.5%) | 2671 (21.3%) |
| 5 (least deprived) | 2700 (25.1%) | 3164 (25.2%) |
| Missing | 1263 (11.7%) | 1453 (11.6%) |
| **Social mixing** |  |  |
| None or all remote | 2685 (25.0%) | 3165 (25.2%) |
| Face to face (with SD) | 6584 (61.4%) | 7647 (61.0%) |
| Physical contact | 1460 (13.6%) | 1675 (13.4%) |
| Missing | 43 (0.4%) | 54 (0.4%) |
| **Comorbidities** |  |  |
| Not diabetic | 9918 (92.1%) | 11,518 (91.8%) |
| Diabetic | 400 (3.9%) | 479 (3.8%) |
| Missing | 454 (4.2%) | 544 (4.3%) |
| **Comorbidities** |  |  |
| Not Immunosuppressed | 9983 (92.7%) | 11,577 (92.3%) |
| Immunosuppressed | 335 (3.1%) | 420 (3.4%) |
| Missing | 454 (4.2%) | 544 (4.3%) |
| **Shielding status** |  |  |
| Not advised to shield | 10,324 (95.8%) | 11,936 (95.2%) |
| Advised to shield | 410 (3.8%) | 554 (4.4%) |
| Missing | 38 (0.4%) | 51 (0.4%) |
| **Smoking status** |  |  |
| Never/ex-smoker | 10,139 (94.1%) | 11,815 (94.2%) |
| Current smoker | 533 (5.0%) | 609 (4.9%) |
| Missing | 533 (5.0%) | 117 (0.9%) |
| **Occupational factors** |  |  |
| **Occupation** |  |  |
| Doctor or medical support | 2596 (24.1%) | - |
| Nurse, NA or Midwife | 2354 (21.9%) | - |
| Allied Health Professional* | 4422 (41.1%) | - |
| Dental | 418 (3.9%) | - |
| Admin, estates or other | 607 (5.6%) | - |
| Missing | 375 (3.5%) | - |
| **Method of commuting** |  |  |
| Alone or with members of household | 9577 (88.9%) | - |
| With others outside household | 1061 (9.9%) | - |
| Missing | 134 (1.2%) | - |
| **Number of SARS-CoV-2 positive patients attended to per week (with physical contact)** |  |  |
| None | 6298 (58.5%) | - |
| 1 – 5 | 2169 (20.1%) | - |
| 6 – 20 | 1506 (14.0%) | - |
| ≥ 21 | 687 (6.4%) | - |
| Missing | 112 (1.0%) | - |
| **Access to appropriate PPE** |  |  |
| Not applicable or all/most of the time | 4560 (42.3%) | - |
| Some of the time or less frequently | 6182 (57.4%) | - |
| Missing | 30 (0.3%) | - |
| **Aerosol generating procedure exposure** |  |  |
| Less than weekly exposure | 8437 (78.3%) | - |
| At least weekly exposure | 2296 (21.3%) | - |
| Missing | 39 (0.4%) | - |
| **Night shift pattern** |  |  |
| Never works nights | 7543 (70.0%) | - |
| Works nights less than weekly | 1796 (16.7%) | - |
| Works nights weekly or always | 1317 (12.2%) | - |
| Missing | 116 (1.1%) | - |
| **Work areas** |  |  |
| Ambulance | 396 (3.7%) | - |
| Community clinical setting / primary care | 2426 (22.5%) | - |
| Non clinical community setting | 565 (5.3%) | - |
| Emergency Department | 963 (8.9%) | - |
| Intensive Care Unit | 927 (8.6%) | - |
| Hospital Inpatient | 2759 (25.6%) | - |
| Hospital Outpatient | 1831 (17.0%) | - |
| Hospital non-clinical area or laboratory | 1114 (10.3%) | - |
| Psychiatric hospital | 312 (2.9%) | - |
| Maternity | 344 (3.2%) | - |
| Nursing or Care Home | 242 (2.3%) | - |
| University | 220 (2.0%) | - |
| Home | 1715 (15.9%) | - |
| Missing (range) † | 34 – 40 (0.3 – 0.4%) | - |
| **Work region** |  |  |
| London | 1423 (13.2%) | - |
| South East England | 1265 (11.7%) | - |
| South West England | 857 (8.0%) | - |
| East of England | 759 (7.1%) | - |
| East Midlands | 1097 (10.2%) | - |
| West Midlands | 834 (7.7%) | - |
| North East England | 445 (4.1%) | - |
| North West England | 1101 (10.2%) | - |
| Yorkshire and the Humber | 778 (7.2%) | - |
| Wales | 334 (3.1%) | - |
| Scotland | 626 (5.8%) | - |
| Northern Ireland | 130 (1.2%) | - |
| Missing | 1123 (10.4%) | - |

\* Also includes pharmacists, healthcare scientists, ambulance workers and those in optical roles.
† When asked about work areas participants could select multiple answers, therefore the work areas variables are 'dummy' variables comparing all those that did not select an area (reference) with all those that did. Given the similar amount of missing data for each of these dummy variables we present a range of number of missing items and proportions.
All occupational factors (other than region of workplace) relate to work circumstances during the weeks following the first UK national lockdown on March 23<sup>rd</sup> 2020.

A description of the cohort who were working during lockdown, stratified by ethnicity, is shown in Supplementary Table 3. Almost all of the predictor variables significantly differed by ethnicity. Age was significantly different by ethnic group (p<0.001), being lower in the Black and Asian cohorts compared to the White cohort (Black 43.5 [IQR 34.5 – 54], Asian 42 [IQR 33 – 51], White 46 [36 – 55]). A greater proportion of Black HCWs lived in areas corresponding to lower IMD quintiles than White HCWs. Religiosity was also significantly different by ethnic group (p<0.001) with much greater proportions of Black and Asian HCWs describing their religion as being extremely important to their everyday lives compared to the White cohort (41.9% [Black], 19.5%[Asian] vs 5.9% [White]). Ethnic distribution was not equal across regions of the UK (p<0.001) with a higher proportion of Black and Asian HCWs practicing in London (26.4% [Black], 21.3% [Asian] vs 11.8% [White]), and a lower proportion practicing in Scotland (2.5% [Black], 4.6% [Asian] vs 7.2% [White]) and South West England (4.2% [Black], 5.2% [Asian] vs 10.1% [White]). 62.0% of White HCWs did not have physical contact with COVID-19 patients, this compares to 49.1% in Asian and 49.6% in Black HCWs.

### Univariable analysis of predictors of SARS-CoV-2 infection

#### Demographic and household predictors

Overall 2,496 (23.2%) of the 10,772 HCWs who worked during lockdown reported evidence of previous infection. Compared to the uninfected participants, the infected participants were younger, with a greater proportion of Black HCWs compared to White HCWs (OR 1.40, 95%CI 1.14 – 1.73, p=0.001) (Table 2). Comparable patterns of association were seen when including HCWs who reported they were not working during lockdown (Supplementary Table 4).

**Table 2.** Description of the cohort working during lockdown stratified by SARS-CoV-2 infection status with unadjusted odds ratios for the association of predictor variables with infection.

|  | Excluding those not working during lockdown |  |  |  |
| --- | --- | --- | --- | --- |
| Variable | Not infected<br>8276 (76.8%) | Infected<br>2496 (23.2%) | Unadjusted OR<br>(95% CI) | P value |
| **Demographic and household factors** |  |  |  |  |
| **Age, med(IQR)** | 46 (36 – 54) | 43 (32 – 52) | 0.83 (0.80 – 0.86) | <0.001 |
| **Sex** |  |  |  |  |
| Male | 2023 (24.5%) | 637 (25.6%) | Ref | - |
| Female | 6238 (75.5%) | 1851 (74.4%) | 0.94 (0.85 – 1.04) | 0.25 |
| **Ethnicity** |  |  |  |  |
| White | 5872 (71.0%) | 1711 (68.6%) | Ref | - |
| Asian | 1555 (18.8%) | 502 (20.1%) | 1.11 (0.99 – 1.24) | 0.08 |
| Black | 328 (4.0%) | 134 (5.4%) | 1.40 (1.14 – 1.73) | 0.001 |
| Mixed | 351 (4.2%) | 95 (3.8%) | 0.93 (0.74 – 1.17) | 0.54 |
| Other | 170 (2.1%) | 54 (2.2%) | 1.09 (0.80 – 1.49) | 0.59 |
| **Migration status** |  |  |  |  |
| Born in UK | 6126 (74.2%) | 1775 (71.2%) | Ref | - |
| Born abroad | 2129 (25.8%) | 718 (28.8%) | 1.16 (1.05 – 1.29) | 0.003 |
| **Religiosity** |  |  |  |  |
| Not religious or not important | 4733 (58.4%) | 1352 (55.4%) | Ref | - |
| Fairly important | 1745 (21.6%) | 523 (21.4%) | 1.05 (0.94 – 1.18) | 0.40 |
| Very important | 806 (10.0%) | 258 (10.6%) | 1.12 (0.96 – 1.30) | 0.16 |
| Extremely important | 815 (10.1%) | 309 (12.7%) | 1.33 (1.15 – 1.53) | <0.001 |
| **Household size, med (IQR)** | 2 (1 – 3) | 2 (1 – 3) | 1.05 (1.02 – 1.08) | 0.004 |
| **Cohabitation** |  |  |  |  |
| Does not live with other key workers | 4401 (53.5%) | 1170 (47.1%) | Ref | - |
| Lives with other key workers | 3830 (46.5%) | 1315 (52.9%) | 1.29 (1.18 – 1.41) | <0.001 |
| **Accommodation** |  |  |  |  |
| Does not have shared spaces | 6808 (82.7%) | 1999 (80.5%) | Ref | - |
| Has shared spaces | 1420 (17.3%) | 485 (19.5%) | 1.16 (1.04 – 1.30) | 0.01 |
| **Index of multiple deprivation quintile** |  |  |  |  |
| 1 (most deprived) | 694 (9.6%) | 262 (11.5%) | 1.23 (1.01 – 1.47) | 0.03 |
| 2 | 1174 (16.3%) | 423 (18.5%) | 1.17 (1.01 – 1.36) | 0.04 |
| 3 | 1491 (20.6%) | 453 (19.8%) | Ref | - |
| 4 | 1777 (24.6%) | 535 (23.4%) | 0.99 (0.85 – 1.14) | 0.86 |
| 5 (least deprived) | 2089 (28.9%) | 611 (26.8%) | 0.95 (0.82 – 1.09) | 0.45 |
| **Social mixing** |  |  |  |  |
| None or all remote | 2005 (24.3%) | 680 (27.4%) | Ref | - |
| Face to face (with SD) | 5155 (62.5%) | 1429 (57.5%) | 0.82 (0.74 – 0.91) | <0.001 |
| Physical contact | 1084 (13.2%) | 376 (15.1%) | 1.02 (0.88 – 1.18) | 0.77 |
| **Comorbidities** |  |  |  |  |
| Not diabetic | 7622 (96.1%) | 2296 (96.2%) | Ref | - |
| Diabetic | 310 (3.9%) | 90 (3.8%) | 0.95 (0.75 – 1.21) | 0.68 |
| **Comorbidities** |  |  |  |  |
| Not immunosuppressed | 7659 (96.6%) | 2324 (97.4%) | Ref | - |
| Immunosuppressed | 273 (3.4%) | 62 (2.6%) | 0.76 (0.57 – 1.01) | 0.06 |
| **Shielding status** |  |  |  |  |
| Not advised to shield | 7917 (96.0%) | 2407 (96.9%) | Ref | - |
| Advised to shield | 332 (4.0%) | 78 (3.1%) | 0.78 (0.60 – 1.00) | 0.05 |
| **Smoking status** |  |  |  |  |
| Never/ex-smoker | 7760 (94.6%) | 2379 (96.3%) | Ref | - |
| Current smoker | 441 (5.4%) | 92 (3.7%) | 0.68 (0.54 – 0.85) | 0.001 |
| **Occupational factors** |  |  |  |  |
| **Occupation** |  |  |  |  |
| Doctor or medical support | 1966 (24.6%) | 630 (26.1%) | Ref | - |
| Nurse, NA or Midwife | 1721 (21.6%) | 633 (26.3%) | 1.14 (1.01 – 1.30) | 0.04 |
| Allied Health Professional* | 3443 (43.1%) | 979 (40.6%) | 0.88 (0.79 – 0.99) | 0.03 |
| Dental | 359 (4.5%) | 59 (2.5%) | 0.51 (0.38 – 0.68) | <0.001 |
| Admin, estates or other | 497 (6.2%) | 110 (4.6%) | 0.68 (0.55 – 0.86) | 0.001 |
| **Method of commuting** |  |  |  |  |
| Alone or with members of household | 7425 (90.9%) | 2152 (87.3%) | Ref | - |
| With others outside household | 748 (9.2%) | 313 (12.7%) | 1.45 (1.26 – 1.66) | <0.001 |
| **Number of SARS-CoV-2 positive patients attended to per week (with physical contact)** |  |  |  |  |
| None | 5268 (64.3%) | 1030 (41.7%) | Ref | - |
| 1 – 5 | 1537 (18.8%) | 632 (25.6%) | 2.10 (1.87 – 2.35) | <0.001 |
| 6 – 20 | 971 (11.9%) | 535 (21.6%) | 2.82 (2.49 – 3.20) | <0.001 |
| ≥ 21 | 412 (5.0%) | 275 (11.1%) | 3.42 (2.90 – 4.04) | <0.001 |
| **Access to appropriate PPE** |  |  |  |  |
| Not applicable or all/most the time | 3701 (44.9%) | 859 (34.5%) | Ref | - |
| Some of the time or less frequently | 4548 (55.1%) | 1634 (65.5%) | 1.55 (1.41 – 1.70) | <0.001 |
| **Aerosol generating procedure exposure** |  |  |  |  |
| Less than weekly exposure | 6613 (80.2%) | 1824 (73.3%) | Ref | - |
| At least weekly exposure | 1631 (19.8%) | 665 (26.7%) | 1.48 (1.33 – 1.64) | <0.001 |
| **Night shift pattern** |  |  |  |  |
| Never works nights | 6013 (73.4%) | 1530 (62.0%) | Ref | - |
| Works nights less than weekly | 1230 (15.0%) | 566 (22.9%) | 1.81 (1.60 – 2.02) | <0.001 |
| Works nights weekly or always | 946 (11.6%) | 371 (15.0%) | 1.54 (1.35 – 1.76) | <0.001 |
| **Work areas†** |  |  |  |  |
| Ambulance | 241 (2.9%) | 155 (6.2%) | 2.20 (1.79 – 2.71) | <0.001 |
| Community clinical setting / primary care | 1983 (24.0%) | 443 (17.8%) | 0.68 (0.61 – 0.76) | <0.001 |
| Non clinical community setting | 465 (5.6%) | 100 (4.0%) | 0.70 (0.56 – 0.87) | 0.001 |
| Emergency Department | 644 (7.8%) | 319 (12.8%) | 1.73 (1.50 – 2.00) | <0.001 |
| Intensive Care Unit | 677 (8.2%) | 250 (10.0%) | 1.25 (1.08 – 1.46) | 0.004 |
| Hospital Inpatient | 1851 (22.5%) | 908 (36.5%) | 1.98 (1.80 – 2.19) | <0.001 |
| Hospital Outpatient | 1404 (17.0%) | 427 (17.2%) | 1.01 (0.90 – 1.14) | 0.89 |
| Hospital non-clinical area or laboratory | 918 (11.1%) | 196 (7.9%) | 0.68 (0.58 – 0.80) | <0.001 |
| Psychiatric hospital | 224 (2.7%) | 88 (3.5%) | 1.31 (1.02 – 1.68) | 0.04 |
| Maternity | 278 (3.4%) | 66 (2.7%) | 0.78 (0.59 – 1.03) | 0.08 |
| Nursing or Care Home | 173 (2.1%) | 69 (2.8%) | 1.33 (1.00 – 1.76) | 0.05 |
| University | 177 (2.2%) | 43 (1.7%) | 0.80 (0.57 – 1.12) | 0.20 |
| Home | 1444 (17.5%) | 271 (10.9%) | 0.57 (0.50 – 0.66) | <0.001 |
| **Work region** |  |  |  |  |
| West Midlands | 627 (8.4%) | 207 (9.4%) | Ref | - |
| London | 1037 (14.0%) | 386 (17.4%) | 1.14 (0.95 – 1.39) | 0.17 |
| South East England | 990 (13.3%) | 275 (12.4%) | 0.86 (0.70 – 1.05) | 0.13 |
| South West England (& Channel Islands) | 723 (9.7%) | 134 (6.1%) | 0.57 (0.45 – 0.73) | <0.001 |
| East of England | 599 (8.1%) | 160 (7.2%) | 0.83 (0.66 – 1.04) | 0.10 |
| East Midlands | 869 (11.7%) | 228 (10.3%) | 0.80 (0.65 – 0.99) | 0.04 |
| North East England | 342 (4.6%) | 103 (4.7%) | 0.91 (0.69 – 1.18) | 0.47 |
| North West England (& Isle of Man) | 773 (10.4%) | 328 (14.8%) | 1.29 (1.06 – 1.58) | 0.01 |
| Yorkshire and the Humber | 569 (7.7%) | 209 (9.4%) | 1.11 (0.89 – 1.39) | 0.35 |
| Wales | 246 (3.3%) | 88 (4.0%) | 1.10 (0.83 – 1.46) | 0.49 |
| Scotland | 549 (7.4%) | 77 (3.5%) | 0.43 (0.32 – 0.57) | <0.001 |
| Northern Ireland | 112 (1.5%) | 18 (0.8%) | 0.52 (0.31 – 0.87) | 0.03 |

Table 2 shows the cohort who worked during lockdown stratified by SARS-CoV-2 infection status, and unadjusted odds ratios for the association of predictor variables with SARS-CoV-2 infection.
\* Also includes pharmacists, healthcare scientists, ambulance workers and those in optical roles.
† When asked about work areas participants could select multiple answers, therefore the work areas variables are ‘dummy’ variables comparing all those that did not select an area (reference) with all those that did. Here we only show the number and proportion of infected/non-infected participants who did select this area. Percentages are computed column-wise other than the total of infected and non-infected HCWs which are computed row-wise.
All occupational factors (other than region of workplace) relate to work circumstances during the weeks following the first UK national lockdown on March 23<sup>rd</sup> 2020.
95%CI – 95% confidence interval, OR – odds ratio, PPE – personal protective equipment, Ref – reference category for categorical variables, SARS-CoV-2 – severe acute respiratory syndrome coronavirus-

#### Occupational predictors

The proportion of HCWs with a reported history of COVID-19 infection was proportionate to the number of patients with confirmed/suspected COVID-19 a HCW attended to (with physical contact). 16.4% of those that had no physical contact with COVID-19 patients were infected, compared to 40.0% of those that attended to ≥21 COVID-19 patients per week (Table 2).

### Multivariable analysis of predictors of SARS-CoV-2 infection

#### Demographic and household predictors

In the working cohort, older HCWs were less likely to be infected (aOR 0.92, 95%CI 0.87 – 0.96, p<0.001 for each decade increase in age). HCWs that lived with other key workers, compared to those that did not, had a small increase in odds of infection (1.17, 1.06 – 1.29, p=0.002). Those who described their religion as extremely important were more likely to report infection than those to whom religion was not important or were not religious (1.28, 1.09 – 1.53, p=0.003). Significant demographic and household predictors were unchanged if those not working during lockdown were included (Table 3).

**Table 3.** Multivariable analysis of factors associated with SARS-CoV-2 infection.

|  | Adjusted for demographic, home and work factors during lockdown (n=10,772) |  | Adjusted for demographic and home factors (n=12,541) |  |
| --- | --- | --- | --- | --- |
| Variable | aOR (95% CI) | p value | aOR (95% CI) | p value |
| **Demographic and household factors** |  |  |  |  |
| **Age\*** | 0.92 (0.87 – 0.96) | <0.001 | 0.85 (0.82 – 0.88) | <0.001 |
| **Sex** |  |  |  |  |
| Male | Ref | - | Ref | - |
| Female | 1.02 (0.91 – 1.15) | 0.74 | 0.91 (0.82 – 1.01) | 0.07 |
| **Ethnicity** |  |  |  |  |
| White | Ref | - | Ref | - |
| Asian | 0.86 (0.74 – 1.00) | 0.05 | 0.83 (0.73 – 0.94) | 0.004 |
| Black | 1.00 (0.78 – 1.27) | 0.96 | 0.93 (0.74 – 1.16) | 0.52 |
| Mixed | 0.84 (0.65 – 1.07) | 0.16 | 0.85 (0.68 – 1.06) | 0.14 |
| Other | 0.80 (0.57 – 1.14) | 0.22 | 0.82 (0.61 – 1.12) | 0.22 |
| **Migration status** |  |  |  |  |
| Born in UK | Ref | - | Ref | - |
| Born abroad | 1.10 (0.97 – 1.25) | 0.13 | 1.09 (0.97 – 1.22) | 0.13 |
| **Religiosity** |  |  |  |  |
| Not important or not religious | Ref | - | Ref | - |
| Fairly important | 1.08 (0.95 – 1.22) | 0.22 | 1.10 (0.98 – 1.23) | 0.11 |
| Very important | 1.05 (0.89 – 1.24) | 0.55 | 1.18 (1.01 – 1.38) | 0.04 |
| Extremely important | 1.29 (1.09 – 1.53) | 0.003 | 1.30 (1.12 – 1.51) | <0.001 |
| **Index of multiple deprivation** |  |  |  |  |
| 1 (most deprived) | 1.02 (0.85 – 1.24) | 0.81 | 1.17 (0.99 – 1.39) | 0.07 |
| 2 | 1.07 (0.92 – 1.26) | 0.37 | 1.11 (0.94 – 1.30) | 0.21 |
| 3 | Ref | - | Ref | - |
| 4 | 1.01 (0.87 – 1.18) | 0.86 | 0.99 (0.86 – 1.15) | 0.94 |
| 5 (least deprived) | 1.04 (0.90 – 1.21) | 0.58 | 1.01 (0.88 – 1.15) | 0.93 |
| **Household size** | 1.02 (0.98 – 1.06) | 0.33 | 1.01 (0.97 – 1.04) | 0.65 |
| **Cohabitation** |  |  |  |  |
| Does not live with other key workers | Ref | - | Ref | - |
| Lives with other key workers | 1.17 (1.06 – 1.29) | 0.002 | 1.27 (1.16 – 1.39) | <0.001 |
| **Accommodation** |  |  |  |  |
| Does not have shared spaces | Ref | - | Ref | - |
| Has shared spaces | 0.93 (0.82 – 1.06) | 0.28 | 1.01 (0.89 – 1.13) | 0.92 |
| **Social mixing with others outside household** |  |  |  |  |
| None / remote only | Ref | - | Ref | - |
| Face to face with social distancing | 0.90 (0.80 – 1.01) | 0.07 | 0.90 (0.81 – 0.99) | 0.04 |
| With physical contact | 0.99 (0.85 – 1.16) | 0.95 | 1.05 (0.90 – 1.20) | 0.56 |
| **Comorbidities** |  |  |  |  |
| Diabetes | 1.08 (0.84 – 1.40) | 0.55 | 1.12 (0.89 – 1.41) | 0.32 |
| Immunosuppression | 1.00 (0.73 – 1.39) | 0.98 | 0.91 (0.69 – 1.21) | 0.52 |
| **Shielding status** |  |  |  |  |
| Not advised to shield | Ref | - | Ref | - |
| Advised to shield | 0.86 (0.64 – 1.15) | 0.31 | 0.81 (0.63 – 1.03) | 0.09 |
| **Smoking status** |  |  |  |  |
| Ex or non-smoker | Ref | - | Ref | - |
| Current smoker | 0.56 (0.44 – 0.72) | <0.001 | 0.63 (0.51 – 0.79) | <0.001 |
| **Region of workplace** |  |  |  |  |
| West Midlands | Ref | - | Ref | - |
| London | 1.15 (0.93 – 1.41) | 0.20 | 1.16 (0.96 – 1.40) | 0.13 |
| South East England | 0.85 (0.69 – 1.06) | 0.14 | 0.90 (0.74 – 1.10) | 0.29 |
| South West England or Channel Islands | 0.59 (0.46 – 0.76) | <0.001 | 0.62 (0.50 – 0.77) | <0.001 |
| East of England | 0.81 (0.64 – 1.02) | 0.08 | 0.82 (0.66 – 1.03) | 0.09 |
| East Midlands | 0.86 (0.69 – 1.07) | 0.18 | 0.85 (0.69 – 1.04) | 0.11 |
| North East England | 0.88 (0.66 – 1.16) | 0.35 | 0.95 (0.73 – 1.23) | 0.70 |
| North West England or Isle of Man | 1.22 (0.99 – 1.51) | 0.07 | 1.25 (1.03 – 1.51) | 0.02 |
| Yorkshire and the Humber | 1.16 (0.92 – 1.47) | 0.20 | 1.11 (0.89 – 1.39) | 0.34 |
| Wales | 1.14 (0.85 – 1.53) | 0.38 | 1.13 (0.87 – 1.48) | 0.37 |
| Scotland | 0.42 (0.31 – 0.56) | <0.001 | 0.48 (0.36 – 0.63) | <0.001 |
| Northern Ireland | 0.51 (0.30 – 0.87) | 0.01 | 0.51 (0.32 – 0.84) | 0.008 |
| **Time between questionnaire rollout and questionnaire completion (per day)** | 1.00 (1.00 – 1.00) | 0.54 | 1.00 (1.00 – 1.00) | 0.35 |
| **Occupational factors** |  |  |  |  |
| **Occupation** |  |  |  |  |
| Doctor or medical support | Ref | - | - | - |
| Nurse, nursing associate or Midwife | 1.35 (1.15 – 1.58) | <0.001 | - | - |
| Allied health professional† | 1.07 (0.92 – 1.23) | 0.40 | - | - |
| Dental | 0.83 (0.61 – 1.13) | 0.24 | - | - |
| Admin, estates or other | 1.17 (0.90 – 1.51) | 0.24 | - | - |
| **Transport to work** |  |  |  |  |
| Alone or with members of household | Ref | - | - | - |
| With others outside household | 1.10 (0.94 – 1.29) | 0.23 | - | - |
| **Number of SARS-CoV-2 positive patients attended to per week (with physical contact)** |  |  |  |  |
| None | Ref | - | - | - |
| 1 – 5 | 1.66 (1.45 – 1.89) | <0.001 | - | - |
| 6 – 20 | 2.09 (1.79 – 2.44) | <0.001 | - | - |
| ≥ 21 | 2.51 (2.05 – 3.08) | <0.001 | - | - |
| **Access to appropriate PPE** |  |  |  |  |
| Not applicable or all/most of the time | Ref | - | - | - |
| Some of the time or less frequently | 1.27 (1.15 – 1.40) | <0.001 | - | - |
| **Aerosol generating procedure exposure** |  |  |  |  |
| Less than weekly exposure | Ref | - | - | - |
| At least weekly exposure | 0.90 (0.79 – 1.03) | 0.12 | - | - |
| **Night shift pattern** |  |  |  |  |
| Never works nights | Ref | - | - | - |
| Works nights less than weekly | 1.10 (0.96 – 1.27) | 0.17 | - | - |
| Works nights weekly or always | 0.85 (0.72 – 1.00) | 0.05 | - | - |
| **Work areas** |  |  |  |  |
| Ambulance | 1.94 (1.51 – 2.49) | <0.001 | - | - |
| Community clinical setting /primary care | 0.91 (0.80 – 1.04) | 0.19 | - | - |
| Non clinical community setting | 0.92 (0.73 – 1.17) | 0.51 | - | - |
| Emergency Department | 1.11 (0.94 – 1.31) | 0.21 | - | - |
| Intensive Care Unit | 0.76 (0.63 – 0.90) | 0.002 | - | - |
| Hospital Inpatient | 1.54 (1.36 – 1.73) | <0.001 | - | - |
| Hospital Outpatient | 0.92 (0.80 – 1.05) | 0.20 | - | - |
| Hospital non-clinical area or laboratory | 0.85 (0.72 – 1.02) | 0.08 | - | - |
| Psychiatric hospital | 1.29 (0.99 – 1.69) | 0.06 | - | - |
| Maternity | 0.67 (0.51 – 0.89) | 0.006 | - | - |
| Nursing or Care Home | 1.36 (1.01 – 1.84) | 0.04 | - | - |
| University | 0.94 (0.66 – 1.35) | 0.75 | - | - |
| Home | 0.80 (0.69 – 0.93) | 0.004 | - | - |

Table 3 shows the results of two multivariable logistic regression analyses, one containing the whole analysed cohort, examining the association of demographic and household factors with infection, and the other containing the cohort working during lockdown, additionally adjusted for occupational factors.
\*for each decade increase in age. † Also includes pharmacists, healthcare scientists, ambulance workers and those in optical roles.
Analyses adjusted for all other variables in the table (with the exception of the exclusion of occupational predictors in the right hand columns – as indicated by the lack of results in the relevant sections).
All occupational factors (other than region of workplace) relate to work circumstances during the weeks following the first UK national lockdown on March 23<sup>rd</sup> 2020. When asked about work areas participants could select multiple answers, therefore the work areas variables are 'dummy' variables comparing all those that did not select an area (reference) with all those that did. Region of workplace is included in the analysis of household and demographic factors as a proxy for the participants region of residence.
aOR – adjusted odds ratio, PPE – personal protective equipment, Ref – reference category for categorical variables, SARS-CoV-2 – severe acute respiratory syndrome coronavirus-2

#### Occupational predictors

Compared to doctors, those working in nursing and midwifery roles were more likely to be infected (1.35, 1.15 – 1.58, p<0.001). The odds of infection were higher for HCWs who attended to a higher number of confirmed COVID-19 patients (with physical contact), with those attending to ≥21 patients per week being two and a half times more likely to be infected compared to those who did not attend to any COVID-19 patients. Compared to those who either did not need PPE or reported access to appropriate PPE each time they needed it, those who reported *not* having access to appropriate PPE at all times were more likely to be infected (1.27, 1.15 – 1.40, p<0.001). Working in ambulance (1.94, 1.51 – 2.49, p<0.001) or hospital inpatient (1.54, 1.36 – 1.73, p<0.001) settings were associated with higher odds of infection, whilst working in an Intensive Care Unit (ICU) setting was associated with lower odds of infection (0.76, 0.63 – 0.90, p<0.001), when compared to those not working in these settings. HCWs working in Scotland and South West England were at approximately half the odds of being infected compared to those working in the West Midlands (Table 3).

### Association of ethnicity with SARS-CoV-2 infection risk

In a model adjusted for age and sex there was an increased risk of infection amongst Black HCWs compared to White HCWs (Figure 2). This association appeared to diminish as more variables were added to the model and, after adjustment for all predictors, differences in odds of infection between Black and White ethnic groups had attenuated.

**Figure 2.**
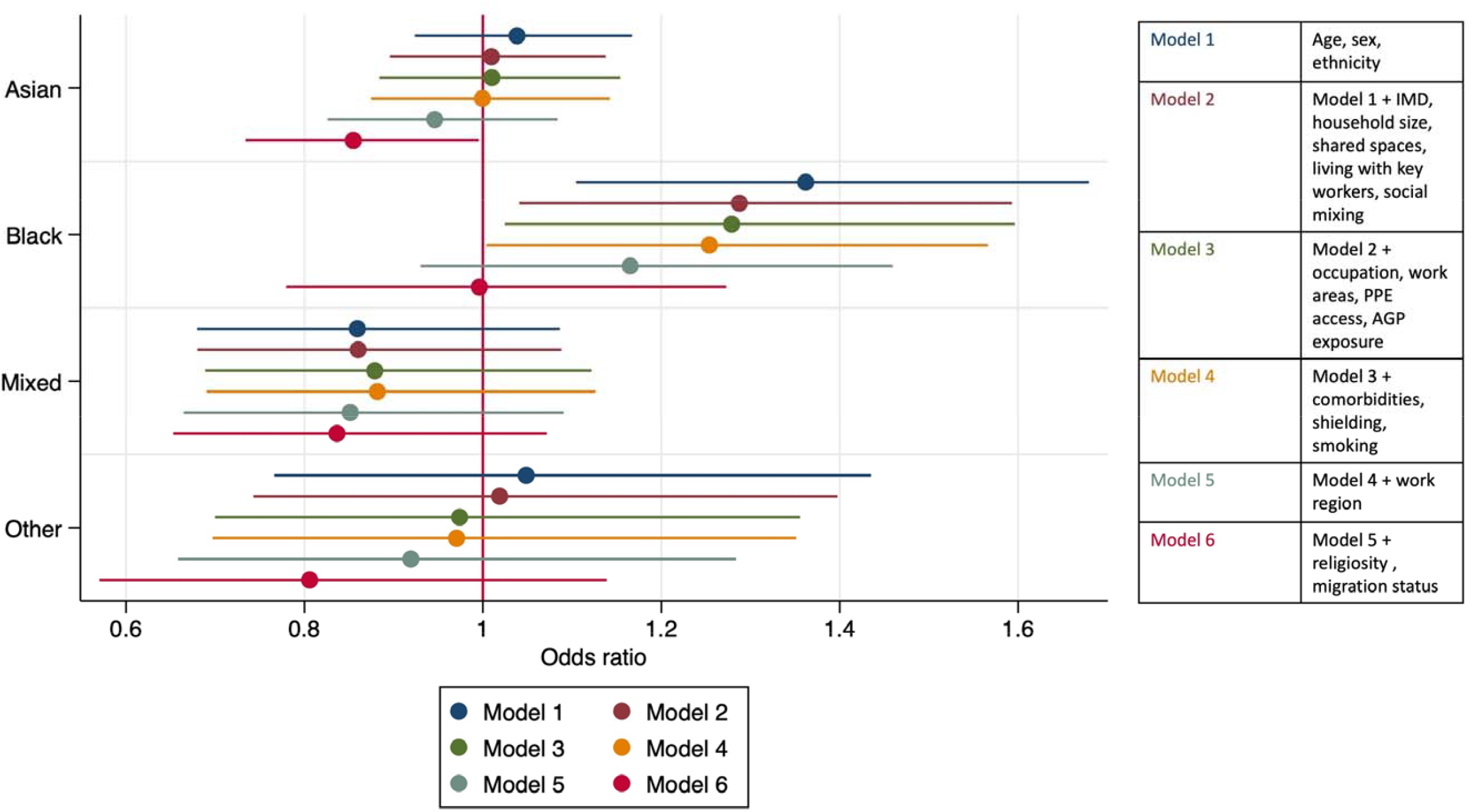
The association of ethnicity and SARS-CoV-2 infection with sequential adjustment for other predictors. Figure 2 shows aORs and 95%CIs for the associations of the five broad ethnic groups (White ethnic group as reference) with SARS-CoV-2 infection and how these changed with sequential adjustment for groups of predictor variables. AGP – aerosol generating procedures; IMD – index of Multiple Deprivation; PPE – personal protective equipment.

### Sensitivity analyses

Results of: i) an analysis using an outcome of infection defined by either positive PCR or antibody and excluding those who had never been tested; ii) an analysis of complete cases; and iii) an analysis investigating the effect of vaccination-induced seropositivity on our results, did not lead to any changes in our interpretation of the data (see supplementary information and supplementary tables 5 and 6).

Univariable and multivariable logistic regression analyses using the more granular ethnicity categories are shown in Supplementary Table 7. In univariable analysis, those from Pakistani and Black African groups were more likely to be infected than their White British colleagues, but as with the main analysis, these effects were attenuated in the fully adjusted model.

## Discussion

In this analysis of over 12,000 UK HCWs, we found that nearly a quarter of participants reported having been infected with SARS-CoV-2 within the first year of the pandemic. The richness of the dataset and ethnic diversity of the cohort has allowed us to identify factors that may explain the disproportionate risks of infection between Black and White HCWs. Additionally, we have identified important predictors of SARS-CoV-2 infection in HCWs, including: working in nursing or midwifery roles, occupational exposure to increasing numbers of patients with COVID-19, lack of access to PPE, cohabiting with another key worker and working in hospital inpatient or ambulance settings. Those working in particular UK regions (Scotland and South West England) had lower odds of infection than those working in the West Midlands, as did those working in ICU settings.

Our estimate of nearly a quarter of HCWs being infected with SARS-CoV-2 aligns with anti-SARS-CoV-2 seroprevalence estimates in UK healthcare settings, which have been reported to range from 10.8 – 44.0%, varying by UK region in which the study was conducted.^4,15-17^ These estimates, including ours, are significantly higher than the estimated seroprevalence in England (prior to the vaccine rollout and after the first wave of the pandemic) which was estimated to be 6%,^18^ adding weight to the suggestion that UK HCWs are at higher risk of SARS-CoV-2 infection than the general population.^18,19^

We demonstrate a strong association between the number of confirmed COVID-19 patients attended to by a HCW and their risk of infection. Previous studies have found conflicting evidence on this point. Caring for COVID-19 patients was found not to increase risk of infection in two large studies conducted in the USA,^3,20^ whereas in the UK, those working in patient-facing roles during the pandemic have been shown to be at higher risk of infection.^4,17^ It should be noted that there are different PPE standards recommended by the two countries (in the USA, HCWs are advised to wear a ‘higher grade’ of PPE)^21^ and this may contribute to the differences in infection risk and the significant predictors of infection for HCWs practising in the two countries.

Evidence for the importance of PPE in preventing HCW infection can be found in our analysis, with those who felt they did not have access to appropriate PPE at all times being more likely to have been infected than those who did. Furthermore, those working in ICU settings (where long sleeve gowns and respirator facemasks are recommended at all times) had lower odds of infection than those working elsewhere. These findings are in agreement with existing studies^4,15,19^ and, together with the mounting evidence for aerosol transmission of SARS-CoV-2,^22,23^ and the suggestion that coughing may generate more aerosols than activities designated as AGPs such as delivery of continuous positive airway pressure^24^ support the claim that upgrading PPE standards for HCWs attending to COVID-19 patients (regardless of whether they are performing AGPs) may have a beneficial impact on the risk of HCW infection.

We found that a higher proportion of those from Black ethnic groups reported having been infected with SARS-CoV-2 compared to their White colleagues. This is commensurate with other studies conducted both in the USA and the UK.^3,4,15,20,25^ Ethnicity is a complex construct; it has been defined as “the social group a person belongs to, and either identifies with or is identified with by others, as a result of a mix of cultural and other factors including language, diet, religion, ancestry and physical features traditionally associated with race”.^26^ Only by a deeper understanding of factors relating to disproportionate SARS-CoV-2 infections in ethnic minority groups compared to White groups can we reduce hospitalisation, intensive care unit admission and death.^27^ One strength of our study comes from the richness of our data, which allows us to determine the contribution that some of these interrelated factors may make to the higher risk of infection faced by HCWs from certain ethnic groups. In the fully adjusted model, there was no significant difference in infection risk between White and Black HCWs suggesting that some covariates included in this model might drive the differences in the odds of infection by ethnicity.

We also found there to be an unequal ethnic distribution across other variables associated with increased odds of infection. For example, a far greater proportion of Black participants, compared to their White colleagues, worked in London and a far smaller proportion worked in Scotland, UK regions with amongst the highest and lowest infection rates respectively. Black HCWs reported far higher religiosity than White HCWs, which was shown to be associated with an increased likelihood of infection in the fully adjusted model. Black HCWs were also more likely to live in areas corresponding to the most deprived quintile and were more likely to have been born abroad than their White colleagues, both factors with a univariable association with higher infection risk. Importantly, our results indicate that sociodemographic and occupational differences between ethnic groups, such as those described above, are likely to be responsible for the increased probability of infection in Black HCWs compared to their White colleagues, as opposed to any innate biological characteristics. These findings have important public health implications given the increased risk of SARS-CoV-2 vaccine hesitancy in UK HCWs from Black ethnic groups.^6^

For the first time, we have found that religiosity (one of the factors interrelated to a person’s ethnicity) is associated with increased odds of SARS-CoV-2 infection. Religion has previously been associated with outcomes from COVID-19 at a population level, with analysis by the ONS showing those from particular religious groups (including Muslims and Hindus) in England and Wales are at higher risk of death from COVID-19 than Christians.^28^ The mechanisms underlying our observation are unclear and warrant further investigation.

Risk of infection differed by UK region of workplace with HCWs in South West England and Scotland being at lower risk than those working in the West Midlands. Both areas have a lower population density and lower proportion of ethnic minorities than the West Midlands,^29-31^ factors associated with a decreased risk of SARS-CoV-2 transmission.^32,33^ Additionally, government imposed restrictions aimed at slowing viral transmission differed between the UK nations and this may have influenced the lower infection risk seen amongst those working in Scotland. ^32^

We found ambulance workers to be at twice the risk of infection compared to those not working in this setting. To our knowledge, we are the first to demonstrate this effect. The reasons underlying this association require further investigation, although may relate to the frontline position of these healthcare workers and their exposure to the most critically unwell COVID-19 patients, with much of this exposure occurring in comparatively uncontrolled settings outside of hospital. In-line with previous work from the UK, we also found nurses/midwives to be at higher risk of infection than those in medical roles.^4^

Increasing age was associated with lower odds of infection. This effect may be due to the close correlation of age with occupational seniority. Senior HCWs spend a greater proportion of their time engaged in managerial and administrative responsibilities, and less time engaged in direct patient care, compared to their junior colleagues. This may lead to less occupational exposure to SARS-CoV-2 and therefore lower infection risk.^34^ Additionally, older HCWs have been shown to report better access to PPE (which may also be related to reduced patient contact compared to junior staff).^35^

Our study has a number of limitations. There was potential for selection/responder bias, however comparison with the NHS workforce shows our sample to be representative, albeit with a lower proportion of ancillary staff (bias in the UK-REACH cohort study has been explored elsewhere^36^). As with any consented cohort study there is the potential for self-selection bias. The cross-sectional nature of the study means we cannot infer the direction of causality, since results may be vulnerable to reverse causation and residual confounding. HCWs who thought they had been infected prior to widespread testing and subsequently tested negative for SARS-CoV-2 infection later in the pandemic would be coded as uninfected in our analysis. We may, therefore, have underestimated infection prevalence. Reassuringly, as noted above, the proportion of infected HCWs is in-line with estimates from other UK studies. PCR and serology status are self-reported, although given the implications of positive SARS-CoV-2 tests in a HCW population, we do not expect recall bias to have much effect on our outcome measure.

In conclusion, we identified key sociodemographic and occupational factors associated with SARS-CoV-2 infection amongst UK HCWs in a large national cohort study. These findings are of urgent public health importance, especially in light of the emergence of a highly transmissible variant of SARS-CoV-2 (omicron), against which vaccination may be less effective.^37^ The results should inform policies aimed at protecting HCWs in future pandemic waves through: individualised risk assessments, proactive vaccination strategies (including the booster vaccines) to those at highest risk and better communication around drivers of infection risk to safeguard the healthcare workforce. Critically, we demonstrate that Black HCWs in the UK are more likely to contract COVID-19 than their White colleagues. We have identified some factors interrelated to ethnicity that may underlie this association. Further work should focus on examining how these factors might mediate any disproportionate infection risk to inform interventions. This is particularly important given the increased prevalence of SARS-CoV-2 vaccine hesitancy in Black HCWs.^14^

## Supporting information

STROBE checklist

## Data Availability

To access data or samples produced by the UK-REACH study, the working group representative must first submit a request to the Core Management Group by contacting the UK-REACH Project Manager in the first instance. For ancillary studies outside of the core deliverables, the Steering Committee will make final decisions once they have been approved by the Core Management Group. Decisions on granting the access to data/materials will be made within eight weeks.
Third party requests from outside the Project will require explicit approval of the Steering Committee once approved by the Core Management Group.
Note that should there be significant numbers of requests to access data and/or samples then a separate Data Access Committee will be convened to appraise requests in the first instance.

## Funding

UK-REACH is supported by a grant from the MRC-UK Research and Innovation (MR/V027549/1) and the Department of Health and Social Care through the National Institute for Health Research (NIHR) rapid response panel to tackle COVID-19. Core funding was also provided by NIHR Biomedical Research Centres. CAM is an NIHR Academic Clinical Fellow (ACF-2018-11-004). DP is supported by the NIHR. KW is funded through an NIHR Career Development Fellowship (CDF-2017-10-008). LBN is supported by an Academy of Medical Sciences Springboard Award (SBF005\1047). ALG was funded by internal fellowships at the University of Leicester from the Wellcome Trust Institutional Strategic Support Fund (204801/Z/16/Z) and the BHF Accelerator Award (AA/18/3/ 34220). MDT holds a Wellcome Trust Investigator Award (WT 202849/Z/ 16/Z) and an NIHR Senior Investigator Award. KK and LJG are supported by the National Institute for Health Research (NIHR) Applied Research Collaboration East Midlands (ARC EM). KK and MP are supported by the NIHR Leicester Biomedical Research Centre (BRC). MP is funded by a NIHR Development and Skills Enhancement Award. This work is carried out with the support of BREATHE-The Health Data Research Hub for Respiratory Health [MC_PC_19004] in partnership with SAIL Databank. BREATHE is funded through the UK Research and Innovation Industrial Strategy Challenge Fund and delivered through Health Data Research UK.

## Data availability statement

To access data or samples produced by the UK-REACH study, the working group representative must first submit a request to the Core Management Group by contacting the UK-REACH Project Manager in the first instance. For ancillary studies outside of the core deliverables, the Steering Committee will make final decisions once they have been approved by the Core Management Group. Decisions on granting the access to data/materials will be made within eight weeks.

Third party requests from outside the Project will require explicit approval of the Steering Committee once approved by the Core Management Group.

Note that should there be significant numbers of requests to access data and/or samples then a separate Data Access Committee will be convened to appraise requests in the first instance.

## Acknowledgements

We would like to thank all the participants who have taken part in this study when the NHS is under immense pressure. We wish to acknowledge the Professional Expert Panel group (Amir Burney, Association of Pakistani Physicians of Northern Europe; Tiffanie Harrison; London North West University Healthcare NHS Trust; Ahmed Hashim, Sudanese Doctors Association; Sandra Kazembe, University Hospitals Leicester NHS Trust; Susie M. Lagrata (Co-chair), Filipino Nurses Association, UK & University College London Hospitals NHS Foundation Trust; Satheesh Mathew, British Association of Physicians of Indian Origin; Juliette Mutuyimana, Kingston Hospitals NHS Trust; Padmasayee Papineni (Co-chair), London North West University Healthcare NHS Trust; Tatiana Monteiro, University Hospitals Leicester NHS Trust), the UK-REACH Stakeholder Group ^38^, the Study Steering Committee, Serco, as well as the following people and organisations for their support in setting up the study from the regulatory bodies: Kerrin Clapton and Andrew Ledgard (General Medical Council), Caroline Kenny (Nursing and Midwifery Council), David Teeman and Lisa Bainbridge (General Dental Council), My Phan and Jenny Clapham (General Pharmaceutical Council), Angharad Jones (General Optical Council), Mark Neale (Pharmaceutical Society of Northern Ireland) and the Health and Care Professions Council.

We would also like to acknowledge the following trusts and sites who recruited participants to the study: Affinity Care, Berkshire Healthcare NHS Trust, Birmingham and Solihull NHS Foundation Trust, Birmingham Community Healthcare NHS Foundation Trust, Black Country Community Healthcare NHS Foundation Trust, Bridgewater Community Healthcare NHS Trust, Central London Community Healthcare NHS Trust, Chesterfield Royal Hospital NHS Foundation Trust, County Durham and Darlington Foundation Trust, Derbyshire Healthcare NHS Foundation Trust, Lancashire Teaching Hospitals NHS Foundation Trust, Lewisham and Greenwich NHS Trust, London Ambulance NHS Trust, Northern Borders, Northumbria Healthcare NHS Foundation Trust, Nottinghamshire Healthcare NHS Foundation Trust, Royal Brompton and Harefield NHS trust, Royal Free NHS Foundation Trust, Sheffield Teaching Hospitals NHS Foundation Trust, South Central Ambulance Service NHS Trust, South Tees NHS Foundation Trust, St George’s University Hospital NHS Foundation Trust, Sussex Community NHS Foundation Trust, University Hospitals Coventry and Warwickshire NHS Trust, University Hospitals of Leicester NHS Trust, University Hospitals Southampton NHS Foundation Trust, Walsall Healthcare NHS Trust and Yeovil District Hospital NHS Foundation Trust.

## Declaration of interest

KK is Director of the University of Leicester Centre for Black Minority Ethnic Health, Trustee of the South Asian Health Foundation and Chair of the Ethnicity Subgroup of the UK Government Scientific Advisory Group for Emergencies (SAGE). SC is Deputy Medical Director of the General Medical Council, UK Honorary Professor, University of Leicester. MP reports grants from Sanofi, grants and personal fees from Gilead Sciences and personal fees from QIAGEN, outside the submitted work.

## Notes

### Clinical Protocols

https://bmjopen.bmj.com/content/bmjopen/11/9/e050647.full.pdf

