## Supplementary information for "Predictors of SARS-CoV-2 infection in a multi-ethnic cohort of United Kingdom healthcare workers: a prospective nationwide cohort study (UK-REACH)"

On behalf of the UK-REACH Study Collaborative Group+

+Manish Pareek (Chief investigator), Laura Gray (University of Leicester), Laura Nellums (University of Nottingham), Anna L Guyatt (University of Leicester), Catherine Johns (University of Leicester), I Chris McManus (University College London), Katherine Woolf (University College London), Ibrahim Akubakar (University College London), Amit Gupta (Oxford University Hospitals), Keith R Abrams (University of York), Martin D Tobin (University of Leicester), Louise Wain (University of Leicester), Sue Carr (University Hospital Leicester), Edward Dove (University of Edinburgh), Kamlesh Khunti (University of Leicester), David Ford (University of Swansea), Robert Free (University of Leicester).

1. Department of Respiratory Sciences, University of Leicester, Leicester, UK

2. Department of Infection and HIV Medicine, University Hospitals of Leicester NHS Trust, Leicester, UK

3. Genetic Epidemiology Research Group, Department of Health Sciences, University of Leicester, Leicester, UK

4. Biostatistics Research Group, Department of Health Sciences, University of Leicester, Leicester, UK

5. Department of Nephrology, University Hospitals of Leicester NHS Trust, Leicester, UK

6. General Medical Council, London, UK

7. Oxford University Hospitals NHS Foundation Trust, Oxford, UK

8. Department of Health Sciences, University of Leicester, Leicester, UK

9. University College London Medical School, London, UK

10. Population and Lifespan Sciences, School of Medicine, University of Nottingham, Nottingham, UK

11. Diabetes Research Centre, University of Leicester, Leicester, UK

12. Centre for Health Economics, University of York, York, UK

**Supplementary information – Effects of vaccination induced seropositivity on results**

Of the 56 participants who declared their antibody date to be later than their vaccination date, and who did not have a positive SARS-CoV-2 PCR test, 43 entered an antibody result later than their questionnaire was completed (and thus it was assumed the antibody date was incorrect). Of the remaining 13, 5 patients suspected that they had had COVID-19. In a multivariable analysis recoding the 8 remaining patients, who could feasibly have had antibodies induced by vaccination rather than infection and would not have been coded as being infected due to PCR or suspected COVID-19, as uninfected results remain unchanged (data not shown).

**Supplementary Table 1. Relative contribution of PCR, serology and suspected COVID-19 to overall number of infections in both cohorts**

| **Condition** | **Number (proportion) meeting condition**  **Excluding those not working during lockdown (n=10,772)** | **Number (proportion) meeting condition**  **Including those not working during lockdown (n=12,541)** |
| --- | --- | --- |
| **PCR positive (negative, not tested or missing for serology)** | 1013 (9.4%) | 1167 (9.3%) |
| **Serology positive (negative, not tested or missing for PCR)** | 899 (8.3%) | 964 (7.7%) |
| **PCR and serology positive** | 412 (3.8%) | 436 (3.5%) |
| **Not tested by PCR or serology (or both missing) but suspected COVID-19** | 172 (1.6%) | 224 (1.8%) |
| **Total infections** | 2496 (23.2%) | 2791 (22.3%) |

COVID-19 – coronavirus disease 2019, n – number, PCR – polymerase chain reaction

**Supplementary Table 2. Derivation of covariates from questionnaire data**

| **Variable** | **Description** |
| --- | --- |
| **Age** | Continuous variable. Age in years. Derived from date of birth entered by participants at registration. |
| **Sex** | Binary variable. Participants were asked their sex assigned at birth. |
| **Ethnicity** | Categorical variable. Participants were asked to select their ethnicity from a list of the 18 Office for National Statistics categories:  Asian/Asian British – Indian  Asian/Asian British – Pakistani  Asian/Asian British – Bangladeshi  Asian/Asian British – Chinese  Asian/Asian British - Any other Asian background  Black/African/Caribbean/Black British - African  Black/African/Caribbean/Black British – Caribbean  Black/African/Caribbean/Black British - Any other Black/African/Caribbean background Mixed/Multiple ethnic groups - White and Black Caribbean  Mixed/Multiple ethnic groups - White and Black African  Mixed/Multiple ethnic groups - White and Asian  Mixed/Multiple ethnic groups - Any other Mixed/multiple ethnic background  White - English/Welsh/Scottish/Northern Irish/British  White – Irish  White - Gypsy or Irish Traveller  White - Any other white background  Other ethnic group – Arab  Other ethnic group - Any other ethnic background  These were categorised into the 5 broader Office for National Statistics ethnicity categories (Asian, Black, Mixed, White, Other). |
| **Migration status** | Binary variable. Participants were asked whether they were born in the UK. |
| **Religiosity** | Ordinal variable. Participants were asked “How important is religion to you in your everyday life?” and could answer using the following scale: not at all important, fairly important, very important, extremely important and prefer not to answer. This question was asked only to those who indicated that they identified as belonging to a particular religious group in a previous question. Those who indicated they had “no religion” were grouped together with those indicating religion was not at all important. |
| **Household size** | Continuous variable. Participants were asked how many people live in their house other than themselves. |
| **Cohabitation (with key workers)** | Binary variable. Participants were asked “Apart from yourself, how many people in your household work in jobs that often bring them into close physical contact (within 2 metres) with others? Some examples include: bus driver, carer, cleaner, doctor, supermarket checkout worker, teacher.”. This was categorised into the binary ‘does not live with another key worker’ vs ‘lives with another key worker’. |
| **Accommodation (contains shared spaces)** | Binary variable. Participants were asked to answer yes or no to the question “Does your accommodation include shared communal areas such as hallways, stairwells or lifts?” |
| **Index of Multiple Deprivation (IMD) quintile** | Ordinal variable. Participants provided their residential postcode on registration for the study. This was used to determine the Index of Multiple Deprivation (the official measure of deprivation for small areas of England) in the area in which they live. The IMD ranks all areas in England based on 7 measures of deprivation and the ranks can be expressed as quintiles. Lower quintiles indicate more deprivation. Although Wales, Scotland and Northern Ireland have their own measures of deprivation, these are said not to be directly comparable to English IMD and therefore we elected to impute an ‘English IMD’ for residents of the these nations. |
| **Social mixing** | Categorical variable. Participants were asked to indicate the number of contacts they had with other outside of their work both face-to-face with social distancing and with physical contact. Answers were used to derive a three level categorical variable: “No social contact or all remote”, “social contact but socially distanced”, “social contact with physical contact”. |
| **Comorbidities (diabetes and immunosuppression)** | Binary variables. Participants were asked to indicate if they had “diabetes (type I or II)” or “ A weakened immune system or reduced ability to deal with infections (as a result of a disease or treatment)” |
| **Shielding status** | Binary variable. Participants were asked “Have you been contacted by letter or text message to say you are at severe risk from COVID-19 due to an underlying health condition and should be shielding?” |
| **Smoking status** | Binary variable. Participants were asked to indicate their current smoking status. Never and ex-smokers were grouped together and compared with current smokers. |
| **Occupation** | Categorical variable. Participants were asked to select their main job/role. Categorised as below:  **Doctor or medical support** - Doctor, Advanced Critical Care Practitioner, Anaesthesia associate, Surgical Care Practitioner, Other medical associate  **Nurse, NA or Midwife -**  Advanced Nurse Practitioner, Healthcare assistant, Maternity support worker, Midwife, Nurse, Nursing Associate, Other nursing and midwifery role,  **Allied Health Professional (including pharmacists, ambulance workers and those in optical roles)** - Arts therapist, Biomedical scientist, Chiropodist/Podiatrist, Clinical scientist, Dietician, Hearing aid dispenser, Occupational therapist, Operating department practitioner, Orthoptist, Physiotherapist, Practitioner psychologist, Prosthetist / Orthotist, Radiographer, Speech and language therapist, Other Allied Health Professional role, Emergency medical , Paramedic , Other ambulance role, OT Support , Phlebotomist, Physiotherapy Assistant, Radiography Other clinical support role , Pharmacist , Pharmacy technician, Other pharmacy role, Optical - Dispensing optician, Optometrist, Other Optical role  **Dental -**  Clinical dental technician, Dental Hygienist, Dental nurse, Dental technician, Dentist, Other dental role  **Admin, estates or other –** Administration, Catering services, Domestic services, Estates services, Porter, Other |
| **Method of commuting** | Binary variable. Participants were asked the following question: “Which of the following modes of transport do you use to commute on a typical working day over the past month? Please select all that apply” and could select from the following options “Car, alone or with member of household, car share, with a small pool of people outside of household, taxi or private hire vehicle, public transport (e.g. bus, train, tram, underground), motorcycle, scooter or moped, bicycle, on foot, other”. Participants who indicated that at least part of their journey contained a means of transportation that was shared with others outside of their household was coded as “with others outside household”. |
| **Number of SARS-CoV-2 positive patients attended to per week (with physical contact)** | Ordinal variable. Participants were asked to select how many suspected or confirmed COVID-19 patients they attended to (with physical contact) in a week on the following scale: 0, 1-5, 6-20, 21-50, 51+. This was categorised as 0, 1-5, 6 – 20, ≥21. |
| **Access to appropriate PPE** | Binary variable. Participants were asked to indicate how frequently they had access to PPE in the first month after the start of UK national lockdown using the following scale “Not applicable, not at all, rarely, some of the time, yes, most of the time, yes, all of the time. This was collapsed into the binary variable “not applicable or all or most of the time” vs “some of the time or less frequently” |
| **Work areas** | Binary (dummy) variables. Participants were asked to select areas in which they work from a list of non-mutually exclusive clinical and non-clinical areas. |
| **Aerosol generating procedure exposure** | Binary variable. Derived from a question that asked how often participants were in a room where aerosol generating procedures are performed. On the following scale: Not applicable, Never, Once a month or less, A few times a month, Once a week, A few times a week, Every day. |
| **Night shift pattern** | Categorical variable. Participants were asked how often they work night shifts. On the following scale: Not applicable, Never, Less than once a month, Once a month or more, but not every week, Once a week or more, but not every shift, I always work nights. This was categorised as never, less than weekly or weekly/always |
| **Work region** | Categorical variable. Participants were asked to enter the first part of the postcode of their place of work. This was mapped to one of 12 UK regions. |

**Supplementary Table 3. Description of the cohort working during lockdown stratified by ethnicity together with tests of association between predictor variables and ethnicity**

| **Variable** | **Whole cohort working during lockdown (n=10,722)** | **White** | **Asian** | **Black** | **Mixed** | **Other** | **P value** |
| --- | --- | --- | --- | --- | --- | --- | --- |
| **Age**, med(IQR)  Missing | 45 (35 – 54)  54 (0.5%) | 46 (36 – 55)  35 (0.5%) | 42 (33 – 51)  12 (0.6%) | 43.5 (34.5 – 54)  2 (0.4%) | 41 (32 – 50)  3 (0.7%) | 43 (35 – 53)  2 (0.9%) | <0.001 |
| **Sex**  Male  Female  Missing | 2660 (24.7%)  8089 (75.1%)  23 (0.2%) | 1485 (19.6%)  6085 (80.3%)  13 (0.2%) | 813 (39.5%)  1241 (60.3%)  3 (0.2%) | 149 (32.3%)  311 (67.3%)  2 (0.4%) | 105 (23.5%)  339 (76.0%)  2 (0.5%) | 108 (48.2%)  113 (50.5%)  3 (1.3) | <0.001 |
| **Migration status**  Born in UK  Born abroad  Missing | 7901 (73.5%)  2847 (26.5%)  24 (0.2%) | 6642 (87.6%)  926 (12.2%)  15 (0.2%) | 734 (35.7%)  1317 (64.0%)  6 (0.3%) | 144 (31.2%)  316 (68.4%)  2 (0.4%) | 340 (76.2%)  106 (23.8%)  0 (0.0%) | 41 (18.3%)  182 (81.3%)  1 (0.5%) | <0.001 |
| **Religiosity**  Not religious /not important  Fairly important  Very important  Extremely important  Missing | 6085 (56.5%)  2268 (21.1%)  1064 (9.9%)  1124 (10.4%)  231 (2.1%) | 5041 (66.5%)  1468 (19.4%)  492 (6.5%)  439 (5.8%)  143 (1.9%) | 624 (30.3%)  613 (29.8%)  373 (18.1%)  390 (19.0%)  57 (2.8%) | 68 (14.7%)  77 (16.7%)  118 (25.5%)  190 (41.1%)  9 (2.0%) | 288 (64.6%)  73 (16.4%)  29 (6.5%)  42 (9.4%)  14 (3.1%) | 64 (28.6%)  37 (16.5%)  52 (23.2%)  63 (28.1%)  8 (3.6%) | <0.001 |
| **Household size,** med (IQR)  Missing | 2 (1 – 3)  8 (0.1%) | 2 (1 – 3)  3 (0.0%) | 2 (1 – 3)  3 (0.2%) | 2 (1 – 3)  0 (0.0%) | 2 (1 – 3)  1 (0.2%) | 2 (1 – 3)  1 (0.5%) | <0.001 |
| **Cohabitation**  Does not live with other key workers  Lives with other key workers  Missing | 5571 (51.7%)  5145 (47.8%)  56 (0.5%) | 4025 (53.1%)  3527 (46.5%)  31 (0.4%) | 970 (47.2%)  1073 (52.2%)  14 (0.7%) | 211 (45.7%)  246 (53.3%)  5 (1.1%) | 246 (55.2%)  197 (44.2%)  3 (0.7%) | 119 (53.1%)  102 (45.5%)  3 (1.3%) | <0.001 |
| **Accommodation**  Does not have shared spaces  Has shared spaces  Missing | 8807 (81.8%)  1905 (17.7%)  60 (0.6%) | 6491 (85.6%)  1054 (13.9%)  38 (0.5%) | 1492 (72.5%)  552 (26.8%)  13 (0.6%) | 327 (70.8%)  133 (28.8%)  2 (0.4%) | 345 (77.4%)  99 (22.2%)  2 (0.5%) | 152 (67.9%)  67 (29.9%)  5 (2.2%) | <0.001 |
| **Index of multiple deprivation quintile**  1 (most deprived)  2  3  4  5 (least deprived)  Missing | 956 (8.9%)  1597 (14.8%)  1944 (18.1%)  2312 (21.5%)  2700 (25.1%)  1263 (11.7%) | 592 (7.8%)  1060 (14.0%)  1375 (18.1%)  1643 (21.7%)  1934 (25.5%)  979 (12.9%) | 199 (9.7%)  337 (16.4%)  339 (16.5%)  477 (23.2%)  523 (25.4%)  182 (8.9%) | 89 (19.3%)  97 (21.0%)  95 (20.6%)  65 (14.1%)  79 (17.1%)  37 (8.0%) | 41 (9.2%)  69 (15.5%)  96 (21.5%)  84 (18.8%)  117 (26.2%)  39 (8,7%) | 35 (15.6%)  34 (15.2%)  39 (17.4%)  43 (19.2%)  47 (21.0%)  26 (11.6%) | <0.001 |
| **Social mixing**  None or all remote  Face to face (with social distancing)  Physical contact  Missing | 2685 (25.0%)  6584 (61.4%)  1460 (13.6%)  43 (0.4%) | 1659 (21.9%)  4874 (64.3%)  1023 (13.5%)  27 (0.4%) | 710 (34.5%)  1095 (53.2%)  242 (11.8%)  10 (0.5%) | 142 (30.7%)  243 (52.5%)  75 (16.2%)  2 (0.4%) | 107 (24.0%)  256 (57.4%)  80 (17.9%)  3 (0.7%) | 67 (29.9%)  116 (51.8%)  40 (17.9%)  1 (0.5%) | <0.001 |
| **Comorbidities**  Not diabetic  Diabetic  Missing | 9918 (92.1%)  400 (3.9%)  454 (4.2%) | 7053 (93.0%)  225 (3.0%)  305 (4.0%) | 1840 (89.5%)  125 (6.1%)  92 (4.5%) | 412 (89.2%)  24 (5.2%)  26 (5.6%) | 411 (92.2%)  18 (4.0%)  17 (3.8%) | 202 (90.2%)  8 (3.6%)  14 (6.3%) | <0.001 |
| **Comorbidities**  Not Immunosuppressed  Immunosuppressed  Missing | 9983 (92.7%)  335 (3.1%)  454 (4.2%) | 7023 (92.6%)  255 (3.4%)  305 (4.0%) | 1917 (93.2%)  48 (2.3%)  92 (4.5%) | 427 (92.4%)  9 (2.0%)  26 (5.6%) | 414 (92.2%)  15 (3.4%)  17 (3.8%) | 202 (90.2%)  8 (3.6%)  14 (6.3%) | 0.09 |
| **Shielding status**  Not advised to shield  Advised to shield  Missing | 10,324 (95.8%)  410 (3.8%)  38 (0.4%) | 7301 (96.3%)  256 (3.4%)  26 (0.3%) | 1960 (95.3%)  89 (4.3%)  8 (0.4%) | 429 (92.9%)  31 (6.7%)  2 (0.4%) | 425 (95.3%)  20 (4.5%)  1 (0.2%) | 209 (93.3%)  14 (6.3%)  1 (0.5%) | 0.008 |
| **Smoking status**  Never/ex-smoker  Current smoker  Missing | 10,139 (94.1%)  533 (5.0%)  533 (5.0%) | 7106 (93.7%)  426 (5.6%)  51 (0.7%) | 1964 (95.5%)  61 (3.0%)  32 (1.6%) | 448 (97.0%)  9 (2.0%)  5 (1.1%) | 409 (91.7%)  28 (6.3%)  9 (2.0%) | 212 (94.6%)  9 (4.0%)  3 (1.3%) | <0.001 |
| **Occupation**  Doctor or medical support  Nurse, NA or Midwife  Allied Health Professional*  Dental  Admin, estates or other  Missing | 2596 (24.1%)  2354 (21.9%)  4422 (41.1%)  418 (3.9%)  607 (5.6%)  375 (3.5%) | 1045 (13.8%)  2026 (26.7%)  3503 (46.2%)  290 (3.8%)  494 (6.5%)  225 (3.0%) | 1092 (53.1%)  159 (7.7%)  550 (26.7%)  96 (4.7%)  70 (3.4%)  90 (4.4%) | 170 (36.8%)  90 (19.5%)  152 (32.9%)  11 (2.4%)  13 (2.8%)  26 (5.6%) | 164 (36.8%)  55 (12.3%)  176 (39.5%)  14 (3.1%)  22 (4.9%)  15 (3.4%) | 125 (55.8%)  24 (10.7%)  41 (18.3%)  7 (3.1%)  8 (3.6%)  19 (8.5%) | <0.001 |
| **Method of commuting**  Alone or with members of household  With others outside household  Missing | 9577 (88.9%)  1061 (9.9%)  134 (1.2%) | 6842 (90.2%)  656 (8.7%)  85 (1.1%) | 1789 (87.0%)  239 (11.6%)  29 (1.4%) | 377 (81.6%)  79 (17.1%)  6 (1.3%) | 390 (87.4%)  48 (10.8%)  8 (1.8%) | 179 (79.9%)  39 (17.4%)  6 (2.7%) | <0.001 |
| **Number of SARS-CoV-2 positive patients attended to per week (with physical contact)**  None  1 – 5  6 – 20  ≥ 21  Missing | 6298 (58.5%)  2169 (20.1%)  1506 (14.0%)  687 (6.4%)  112 (1.0%) | 4702 (62.0%)  960 (12.7%)  440 (5.8%)  162 (7.9%)  68 (0.9%) | 1009 (49.1%)  503 (24.5%)  354 (17.2%)  162 (7.9%)  29 (1.4%) | 229 (49.6%)  118 (25.5%)  78 (16.9%)  30 (6.5%)  7 (1.5%) | 254 (57.0%)  81 (18.2%)  71 (15.9%)  36 (8.1%)  4 (0.9%) | 104 (46.4%)  54 (24.1%)  43 (19.2%)  19 (8.5%)  4 (1.8%) | <0.001 |
| **Access to appropriate PPE**  Not applicable or all/most the time  Some of the time or less frequently  Missing | 4560 (42.3%)  6182 (57.4%)  30 (0.3%) | 3383 (44.6%)  4183 (55.2%)  17 (0.2%) | 713 (34.7%)  1336 (65.0%)  8 (0.4%) | 188 (40.7%)  270 (58.4%)  4 (0.9%) | 186 (41.7%)  259 (58.1%)  1 (0.2%) | 90 (40.2%)  134 (59.8%)  0 (0.0%) | <0.001 |
| **Aerosol generating procedure exposure**  Less than weekly exposure  At least weekly exposure  Missing | 8437 (78.3%)  2296 (21.3%)  39 (0.4%) | 6115 (80.6%)  1449 (19.1%)  19 (0.3%) | 1482 (72.1%)  562 (27.3%)  13 (0.6%) | 342 (74.0%)  115 (24.9%)  5 (1.1%) | 345 (77.4%)  100 (22.4%)  1 (0.2%) | 153 (68.3%)  70 (31.3%)  1 (0.5%) | <0.001 |
| **Night shift pattern**  Never works nights  Works nights less than weekly  Works nights weekly or always  Missing | 7543 (70.0%)  1796 (16.7%)  1317 (12.2%)  116 (1.1%) | 5626 (74.2%)  1090 (14.4%)  799 (10.5%)  68 (0.9%) | 1238 (60.2%)  456 (22.2%)  329 (16.0%)  34 (1.7%) | 271 (58.7%)  100 (21.7%)  81 (17.5%)  10 (2.2%) | 290 (65.0%)  98 (22.0%)  55 (12.3%)  3 (0.7%) | 118 (52.7%)  52 (23.2%)  53 (23.7%)  1 (0.5%) | <0.001 |
| **Work areas**  Ambulance  Community clinical setting / primary care  Non clinical community setting  Emergency Department  Intensive Care Unit  Hospital Inpatient  Hospital Outpatient  Hospital non-clinical area or laboratory  Psychiatric hospital  Maternity  Nursing or Care Home  University  Home  Missing (range) † | 396 (3.7%)  2426 (22.5%)  565 (5.3%)  963 (8.9%)  927 (8.6%)  2759 (25.6%)  1831 (17.0%)  1114 (10.3%)  312 (2.9%)  344 (3.2%)  242 (2.3%)  220 (2.0%)  1715 (15.9%)  34 – 40 (0.3 – 0.4%) | 359 (4.7%)  1731 (22.8%)  462 (6.1%)  536 (7.1%)  619 (8.2%)  1696 (22.4%)  1140 (15.0%)  787 (10.4%)  211 (2.8%)  235 (3.1%)  202 (2.7%)  162 (2.1%)  1249 (16.5%)  19 – 23 (0.3 – 0.3%) | 18 (0.9%)  479 (23.3%)  59 (2.9%)  290 (14.1%)  203 (9.9%)  714 (34.7%)  485 (23.6%)  212 (10.3%)  61 (3.0%)  68 (3.3%)  20 (1.0%)  35 (1.7%)  304 (14.8%)  8 – 11 (0.4 – 0.5%) | $  93 (20.1%)  17 (3.7%)  56 (12.1%)  39 (8.4%)  133 (28.8%)  69 (14.9%)  51 (11.0%)  26 (5.6%)  21 (4.6%)  15 (3.3%)  7 (1.5%)  58 (12.6%)  4 – 5 (0.9 – 1.1%) | 13 (2.9%)  97 (21.8%)  20 (4.5%)  46 (10.3%)  42 (9.4%)  118 (26.5%)  74 (16.6%)  41 (9.2%)  10 (2.2%)  13 (2.9%)  $  8 (1.8%)  72 (16.1%)  0 (0.0%) | $  26 (11.6%)  7 (3.1%)  35 (15.6%)  24 (10.7%)  98 (43.8%)  63 (28.1%)  23 (10.3%)  $  7 (3.1%)  $  8 (3.6%)  32 (14.3%)  2 – 3 (0.9 – 1.3%) | <0.001  0.001  <0.001  <0.001  0.02  <0.001  <0.001  0.31  0.004  0.12  <0.001  0.07  0.04 |
| **Work region**  London  South East England  South West England  East of England  East Midlands  West Midlands  North East England  North West England  Yorkshire and the Humber  Wales  Scotland  Northern Ireland  Missing | 1423 (13.2%)  1265 (11.7%)  857 (8.0%)  759 (7.1%)  1097 (10.2%)  834 (7.7%)  445 (4.1%)  1101 (10.2%)  778 (7.2%)  334 (3.1%)  626 (5.8%)  130 (1.2%)  1123 (10.4%) | 816 (10.8%)  922 (12.2%)  699 (9.2%)  525 (6.9%)  811 (10.7%)  544 (7.2%)  357 (4.7%)  793 (10.5%)  577 (7.6%)  256 (3.4%)  498 (6.6%)  108 (1.4%)  677 (8.9%) | 373 (18.1%)  214 (10.4%)  90 (4.4%)  152 (7.4%)  191 (9.3%)  192 (9.3%)  68 (3.3%)  193 (9.4%)  129 (6.3%)  50 (2.4%)  81 (3.9%)  13 (0.6%)  311 (15.1%) | 107 (23.2%)  44 (9.5%)  17 (3.7%)  38 (8.2%)  44 (9.5%)  48 (10.4%)  $  40 (8.7%)  37 (8.0%)  11 (2.4%)  10 (2.2%)  $  57 (12.3%) | 78 (17.5%)  63 (14.1%)  38 (8.5%)  33 (7.4%)  39 (8.7%)  34 (7.6%)  11 (2.5%)  48 (10.8%)  26 (5.8%)  9 (2.0%)  25 (5.6%)  $  40 (9.0%) | 49 (21.9%)  22 (9.8%)  13 (5.8%)  11 (4.9%)  12 (5.4%)  16 (7.1%)  $  27 (12.0%)  9 (4.0%)  8 (3.6%)  12 (5.4%)  $  38 (17.0%) | <0.001 |

* Also includes pharmacists, healthcare scientists, ambulance workers and those in optical roles.

† Work areas are binary ‘dummy’ variables which compares all those who selected an area (from a non-mutually exclusive list) to those that did not. Therefore there are different amounts of missing data for each dummy variable and this is presented as a range.

Percentages are computed column-wise . P values are from chi-squared tests for categorical variables and Kruskal-Wallis tests for continuous variables.

IQR – interquartile range; med – median; NA – nursing associate; PPE – personal protective equipment;

**Supplementary Table 4. Description of cohort, including those not working during lockdown, by infection status**

| **Variable** | **Not infected**  **9750 (77.7%)** | **Infected**  **2791 (22.3%)** | **Unadjusted OR (95% CI)** | **P value** |
| --- | --- | --- | --- | --- |
| **Age**, med(IQR) | 45 (35 – 55) | 42 (32 – 52) | 0.84 (0.81 – 0.87) | <0.001 |
| **Sex**  Male  Female | 2289 (23.5%)  7441 (76.5%) | 688 (24.7%)  2094 (75.3%) | Ref  0.94 (0.85 – 1.03) | -  0.18 |
| **Ethnicity**  White  Asian  Black  Mixed  Other | 6883 (70.6%)  1856 (19.0%)  391 (4.0%)  417 (4.3%)  203 (2.1%) | 1912 (68.5%)  562 (20.1%)  144 (5.2%)  112 (4.0%)  61 (2.2%) | Ref  1.09 (0.98 – 1.21)  1.33 (1.09 – 1.62)  0.97 (0.78 – 1.20)  1.08 (0.81 – 1.45) | -  0.12  0.005  0.76  0.60 |
| **Migration status**  Born in UK  Born abroad | 7188 (73.9%)  2536 (26.1%) | 1983 (71.1%)  805 (28.9%) | Ref  1.15 (1.05 – 1.26) | -  0.003 |
| **Religiosity**  Not religious  Fairly important  Very important  Extremely important | 5549 (58.3%)  2049 (21.5%)  942 (9.9%)  985 (10.3%) | 1494 (54.8%)  588 (21.6%)  296 (10.9%)  350 (12.8%) | Ref  1.07 (0.96 – 1.19)  1.17 (1.01 – 1.35)  1.31 (1.15 – 1.50) | -  0.22  0.03  <0.001 |
| **Household size,** med (IQR) | 2 (1 – 3) | 2 (1 – 3) | 1.05 (1.02 – 1.08) | 0.002 |
| **Cohabitation**  Does not live with other key workers  Lives with other key workers | 5255 (54.2%)  4433 (45.8%) | 1319 (47.5%)  1459 (52.5%) | Ref  1.31 (1.21 – 1.43) | -  <0.001 |
| **Accommodation**  Does not have shared spaces  Has shared spaces | 8019 (82.7%)  1673 (17.3%) | 2229 (80.3%)  548 (19.7%) | Ref  1.18 (1.06 – 1.31) | -  0.002 |
| **IMD**  1 (most deprived)  2  3  4  5 (least deprived) | 806 (9.4%)  1376 (16.1%)  1796 (21.0%)  2087 (24.4%)  2477 (29.0%) | 306 (12.0%)  464 (18.2%)  505 (19.8%)  584 (22.9%)  687 (27.0%) | 1.32 (1.12 – 1.55)  1.18 (1.01 – 1.38)  Ref  0.99 (0.86 – 1.14)  0.96 (0.84 – 1.10) | 0.001  0.04  -  0.84  0.57 |
| **Social mixing**  None or all remote  Face to face (with SD)  Physical contact | 2402 (24.7%)  6044 (62.3%)  1262 (13.0%) | 763 (27.5%)  1603 (57.7%)  413 (14.9%) | Ref  0.84 (0.76 – 0.92)  1.03 (0.90 – 1.18) | -  <0.001  0.67 |
| **Comorbidities**  Not diabetic  Diabetic | 8963 (96.0%)  373 (4.0%) | 2555 (96.0%)  106 (4.0%) | Ref  1.01 (0.81 – 1.26) | -  0.95 |
| **Comorbidities**  Not immunosuppressed  Immunosuppressed | 8993 (96.3%)  343 (3.7%) | 2584 (97.1%)  77 (2.9%) | Ref  0.78 (0.61 – 1.01) | -  0.06 |
| **Shielding status**  Not advised to shield  Advised to shield | 9260 (95.3%)  454 (4.7%) | 2676 (96.4%)  100 (3.6%) | Ref  0.77 (0.61 – 0.95) | -  0.02 |
| **Smoking status**  Never/ex-smoker  Current smoker | 9152 (94.7%)  510 (5.3%) | 2663 (96.4%)  99 (3.6%) | Ref  0.67 (0.54 – 0.83) | -  <0.001 |
| **Work region**  West Midlands  London  South East England  South West England  East of England  East Midlands  North East England  North West England  Yorkshire and the Humber  Wales  Scotland  Northern Ireland | 743 (8.5%)  1225 (14.0%)  1177 (13.5%)  843 (9.7%)  742 (8.5%)  977 (11.2%)  382 (4.4%)  906 (10.4%)  681 (7.8%)  290 (3.3%)  627 (7.2%)  130 (1.5%) | 230 (9.3%)  432 (17.5%)  308 (12.5%)  156 (6.3%)  183 (7.4%)  256 (10.4%)  113 (4.6%)  353 (14.3%)  233 (9.4%)  99 (4.0%)  91 (3.7%)  20 (0.8%) | Ref  1.16 (0.96 – 1.39)  0.85 (0.70 – 1.04)  0.60 (0.49 – 0.75)  0.80 (0.64 – 0.99)  0.84 (0.69 – 1.03)  0.96 (0.74 – 1.24)  1.26 (1.04 – 1.52)  1.11 (0.89 – 1.38)  1.12 (0.86 – 1.46)  0.47 (0.36 – 0.61)  0.53 (0.33 – 0.85) | -  0.12  0.11  <0.001  0.05  0.09  0.74  0.02  0.35  0.40  <0.001  0.01 |

* Also includes pharmacists, healthcare scientists, ambulance workers and those in optical roles.

Percentages are computed column-wise other than the total of infected and non-infected HCWs which are computed row-wise.

95%CI – 95% confidence interval, OR – odds ratio, Ref – reference category for categorical variables

**Supplementary Table 5. Univariable and multivariable analysis of factors associated with SARS-CoV-2 infection as defined by positive PCR or serology and excluding those never tested (n=9,213)**

| Variable | Univariable | | Multivariable | |
| --- | --- | --- | --- | --- |
|  | OR (95% CI) | p value | aOR (95% CI) | p value |
| **Age*** | 0.84 (0.80 – 0.87) | <0.001 | 0.92 (0.88 – 0.97) | 0.001 |
| **Sex**  Male  Female | Ref  0.92 (0.83 – 1.03) | -  0.15 | Ref  1.01 (0.89 – 1.15) | -  0.82 |
| **Ethnicity**  White  Asian  Black  Mixed  Other | Ref  1.15 (1.02 – 1.30)  1.48 (1.19 – 1.85)  0.96 (0.75 – 1.22)  1.13 (0.82 – 1.56) | -  0.02  <0.001  0.72  0.44 | Ref  0.88 (0.75 – 1.03)  1.02 (0.79 – 1.33)  0.87 (0.67 – 1.12)  0.83 (0.58 – 1.18) | -  0.11  0.85  0.28  0.29 |
| **Migration status**  Born in UK  Born abroad | Ref  1.24 (1.12 – 1.38) | -  <0.001 | Ref  1.15 (1.01 – 1.32) | -  0.04 |
| **Religiosity**  Not important or not religious  Fairly important  Very important  Extremely important | Ref  1.08 (0.96 – 1.22)  1.20 (1.02 – 1.40)  1.34 (1.15 – 1.56) | -  0.21  0.03  <0.001 | Ref  1.10 (0.97 – 1.26)  1.13 (0.94 – 1.35)  1.30 (1.09 – 1.54) | -  0.14  0.20  0.003 |
| **Index of multiple deprivation**  1 (most deprived)  2  3  4  5 (least deprived) | 1.19 (0.99 – 1.42)  1.16 (0.99 – 1.36)  Ref  0.99 (0.86 – 1.15)  0.92 (0.79 – 1.06) | 0.06  0.07  -  0.90  0.25 | 1.16 (0.98 – 1.37)  1.10 (0.95 – 1.28)  Ref  0.99 (0.87 – 1.13)  1.01 (0.89 – 1.15) | 0.08  0.21  -  0.89  0.84 |
| **Household size** | 1.04 (1.01 – 1.08) | 0.02 | 1.01 (0.97 – 1.05) | 0.65 |
| **Cohabitation**  Does not live with other key workers  Lives with other key workers | Ref  1.29 (1.18 – 1.42) | -  <0.001 | Ref  1.19 (1.07 – 1.32) | -  0.001 |
| **Accommodation**  Does not have shared spaces  Has shared spaces | Ref  1.18 (1.05 – 1.33) | -  0.007 | Ref  0.95 (0.83 – 1.09) | -  0.47 |
| **Social mixing with others outside household**  None / remote only  Face to face with social distancing  With physical contact | Ref  0.80 (0.71 – 0.89)  0.99 (0.85 – 1.15) | -  <0.001  0.87 | Ref  0.87 (0.77 – 0.98)  0.99 (0.84 – 1.16) | -  0.02  0.88 |
| **Comorbidities**  Diabetes  Immunosuppression | 0.99 (0.77 – 1.28)  0.71 (0.52 – 0.95) | 0.95  0.02 | 1.13 (0.86 – 1.48)  0.90 (0.64 – 1.27) | 0.38  0.56 |
| **Shielding status**  Not advised to shield  Advised to shield | Ref  0.80 (0.62 – 1.04) | -  0.10 | Ref  0.91 (0.68 – 1.23) | -  0.55 |
| **Smoking status**  Ex or non-smoker  Current smoker | Ref  0.63 (0.50 – 0.81) | -  <0.001 | Ref  0.52 (0.40 – 0.67) | -  <0.001 |
| **Time between questionnaire rollout and questionnaire completion (per day)** | 1.00 (1.00 – 1.00) | 0.69 | 1.00 (1.00 – 1.00) | 0.63 |
| **Occupation**  Doctor or medical support  Nurse, nursing associate or Midwife  Allied health professional†  Dental  Admin, estates or other | Ref  1.10 (0.96 – 1.26)  0.87 (0.77 – 0.98)  0.49 (0.36 – 0.68)  0.66 (0.52 – 0.85) | -  0.16  0.02  <0.001  0.001 | Ref  1.34 (1.13 – 1.59)  1.07 (0.92 – 1.25)  0.80 (0.57 – 1.13)  1.16 (0.87 – 1.53) | -  0.001  0.36  0.21  0.31 |
| **Transport to work**  Alone or with members of household  With others outside household | Ref  1.39 (1.20 – 1.61) | -  <0.001 | Ref  1.08 (0.91 – 1.27) | -  0.38 |
| **Number of SARS-CoV-2 positive patients attended to per week (with physical contact)**  None  1 – 5  6 – 20  ≥ 21 | Ref  2.08 (1.85 – 2.35)  2.79 (2.45 – 3.18)  3.35 (2.82 – 3.98) | -  <0.001  <0.001  <0.001 | Ref  1.65 (1.44 – 1.90)  2.08 (1.77 – 2.44)  2.51 (2.03 – 3.10) | -  <0.001  <0.001  <0.001 |
| **Access to appropriate PPE**  Not applicable or all/most of the time  Some of the time or less frequently | Ref  1.49 (1.35 – 1.64) | -  <0.001 | Ref  1.24 (1.12 – 1.38) | -  <0.001 |
| **Aerosol generating procedure exposure**  Less than weekly exposure  At least weekly exposure | Ref  1.45 (1.30 – 1.62) | -  <0.001 | Ref  0.91 (0.80 – 1.05) | -  0.20 |
| **Night shift pattern**  Never works nights  Works nights less than weekly  Works nights weekly or always | Ref  1.76 (1.56 – 1.98)  1.53 (1.33 – 1.75) | -  <0.001  <0.001 | Ref  1.09 (0.94 – 1.26)  0.85 (0.72 – 1.01) | -  0.26  0.07 |
| **Work areas**  Ambulance  Community clinical setting /primary care  Non clinical community setting  Emergency Department  Intensive Care Unit  Hospital Inpatient  Hospital Outpatient  Hospital non-clinical area or laboratory  Psychiatric hospital  Maternity  Nursing or Care Home  University  Home | 2.10 (1.69 – 2.60)  0.67 (0.60 – 0.76)  0.70 (0.55 – 0.90)  1.70 (1.46 – 1.96)  1.19 (1.02 – 1.39)  1.95 (1.76 – 2.15)  0.97 (0.86 – 1.10)  0.65 (0.55 – 0.77)  1.25 (0.96 – 1.62)  0.78 (0.59 – 1.03)  1.27 (0.95 – 1.70)  0.85 (0.60 – 1.22)  0.57 (0.49 – 0.67) | <0.001  <0.001  0.005  <0.001  0.03  <0.001  0.68  <0.001  0.10  0.08  0.11  0.39  <0.001 | 1.90 (1.47 – 2.47)  0.90 (0.78 – 1.04)  0.94 (0.73 – 1.22)  1.12 (0.94 – 1.32)  0.74 (0.62 – 0.89)  1.53 (1.36 – 1.74)  0.91 (0.79 – 1.05)  0.83 (0.69 – 1.00)  1.29 (0.98 – 1.69)  0.69 (0.52 – 0.93)  1.33 (0.98 – 1.82)  0.97 (0.66 – 1.41)  0.79 (0.67 – 0.93) | <0.001  0.15  0.66  0.20  0.001  <0.001  0.19  0.05  0.07  0.01  0.07  0.86  0.005 |
| **Work region**  West Midlands  London  South East England  South West England  East of England  East Midlands  North East England  North West England  Yorkshire and the Humber  Wales  Scotland  Northern Ireland | Ref  1.12 (0.92 – 1.37)  0.85 (0.68 – 1.07)  0.59 (0.46 – 0.76)  0.88 (0.68 – 1.14)  0.82 (0.66 – 1.03)  0.92 (0.69 – 1.23)  1.34 (1.09 – 1.66)  1.13 (0.90 – 1.42)  1.29 (0.95 – 1.74)  0.51 (0.37 – 0.70)  0.59 (0.31 – 1.07) | -  0.26  0.16  <0.001  0.33  0.09  0.58  0.006  0.29  0.11  <0.001  0.08 | Ref  1.12 (0.90 – 1.39)  0.85 (0.67 – 1.07)  0.61 (0.47 – 0.80)  0.86 (0.65 – 1.13)  0.89 (0.70 – 1.13)  0.90 (0.66 – 1.22)  1.29 (1.03 – 1.61)  1.17 (0.93 – 1.49)  1.37 (0.99 – 1.88)  0.48 (0.35 – 0.67)  0.59 (0.32 – 1.10) | -  0.31  0.17  <0.001  0.29  0.33  0.49  0.02  0.19  0.06  <0.001  0.10 |

Supplementary Table 5 shows the results of univariable and multivariable logistic regression analyses, examining the association of covariates with infection, in the cohort working during lockdown.

*for each decade increase in age. † Also includes pharmacists, healthcare scientists, ambulance workers and those in optical roles.

Analyses adjusted for all other variables in the table.

All occupational factors (other than region of workplace) relate to work circumstances during the weeks following the first UK national lockdown on March 23^rd^ 2020. When asked about work areas participants could select multiple answers , therefore the work areas variables are ‘dummy’ variables comparing all those that did not select an area (reference) with all those that did. Region of workplace is included in the analysis of household and demographic factors as a proxy for the participants region of residence.

aOR – adjusted odds ratio, PPE – personal protective equipment, Ref – reference category for categorical variables, SARS-CoV-2 – severe acute respiratory syndrome coronavirus-2

**Supplementary Table 6. Multivariable analysis of factors associated with SARS-CoV-2 infection in complete cases**

|  | Adjusted for demographic, home and work factors during lockdown (n=5,711) | | Adjusted for demographic and home factors (n=7,094) | |
| --- | --- | --- | --- | --- |
| Variable | aOR (95% CI) | p value | aOR (95% CI) | p value |
| **Demographic and household factors** | | | | |
| **Age*** | 0.94 (0.88 – 1.00) | 0.06 | 0.85 (0.81 – 0.89) | <0.001 |
| **Sex**  Male  Female | Ref  1.04 (0.88 – 1.22) | -  0.63 | Ref  0.91 (0.80 – 1.05) | -  0.19 |
| **Ethnicity**  White  Asian  Black  Mixed  Other | Ref  0.84 (0.74 – 1.00)  1.02 (0.73 – 1.43)  0.83 (0.60 – 1.15)  0.97 (0.60 – 1.59) | -  0.10  0.89  0.26  0.91 | Ref  0.81 (0.68 – 0.96)  0.92 (0.69 – 1.23)  0.84 (0.63 – 1.11)  0.86 (0.57 – 1.29) | -  0.02  0.58  0.21  0.46 |
| **Migration status**  Born in UK  Born abroad | Ref  1.17 (0.98 – 1.39) | -  0.08 | Ref  1.14 (0.98 – 1.32) | -  0.09 |
| **Religiosity**  Not important or not religious  Fairly important  Very important  Extremely important | Ref  1.10 (0.93 – 1.30)  0.97 (0.76 – 1.23)  1.26 (1.01 – 1.58) | -  0.26  0.78  0.04 | Ref  1.07 (0.92 – 1.24)  1.13 (0.92 – 1.38)  1.21 (0.99 – 1.46) | -  0.36  0.24  0.06 |
| **Index of multiple deprivation**  1 (most deprived)  2  3  4  5 (least deprived) | 1.08 (0.84 – 1.38)  1.04 (0.84 – 1.29)  Ref  1.00 (0.83 – 1.22)  1.05 (0.87 – 1.27) | 0.57  0.69  -  0.94  0.59 | 1.34 (1.08 – 1.66)  1.06 (0.88 – 1.29)  Ref  0.99 (0.84 – 1.18)  1.03 (0.88 – 1.22) | 0.07  0.52  -  0.94  0.69 |
| **Household size** | 1.01 (0.96 – 1.06) | 0.70 | 1.02 (0.97 – 1.07) | 0.41 |
| **Cohabitation**  Does not live with other key workers  Lives with other key workers | Ref  1.17 (1.02 – 1.34) | -  0.03 | Ref  1.26 (1.12 – 1.42) | -  <0.001 |
| **Accommodation**  Does not have shared spaces  Has shared spaces | Ref  0.95 (0.79 – 1.14) | -  0.57 | Ref  1.03 (0.88 – 1.20) | -  0.73 |
| **Social mixing with others outside household**  None / remote only  Face to face with social distancing  With physical contact | Ref  0.95 (0.82 – 1.11)  1.04 (0.84 – 1.29) | -  0.07  0.72 | Ref  0.93 (0.81 – 1.06)  1.11 (0.92 – 1.33) | -  0.26  0.28 |
| **Comorbidities**  Diabetes  Immunosuppression | 1.15 (0.81 – 1.62)  0.87 (0.56 – 1.34) | 0.43  0.53 | 1.26 (0.95 – 1.69)  0.76 (0.52 – 1.11) | 0.11  0.15 |
| **Shielding status**  Not advised to shield  Advised to shield | Ref  1.08 (0.73 – 1.60) | -  0.69 | Ref  0.91 (0.65 – 1.26) | -  0.56 |
| **Smoking status**  Ex or non-smoker  Current smoker | Ref  0.61 (0.43 – 0.85) | -  0.004 | Ref  0.59 (0.44 – 0.80) | -  0.001 |
| **Region of workplace†**  West Midlands  London  South East England  South West England or Channel Islands  East of England  East Midlands  North East England  North West England or Isle of Man  Yorkshire and the Humber  Wales, Scotland or Northern Ireland | Ref  1.18 (0.91 – 1.53)  0.89 (0.69 – 1.16)  0.65 (0.48 – 0.88)  0.77 (0.58 – 1.04)  0.96 (0.72 – 1.29)  0.99 (0.71 – 1.40)  1.31 (1.00 – 1.71)  1.28 (0.95 – 1.72)  0.85 (0.36 – 2.05) | -  0.22  0.40  0.005  0.09  0.79  0.98  0.05  0.10  0.72 | Ref  1.14 (0.91 – 1.42)  0.90 (0.71 – 1.13)  0.62 (0.47 – 0.81)  0.80 (0.62 – 1.04)  0.94 (0.72 – 1.21)  0.93 (0.69 – 1.26)  1.30 (1.03 – 1.64)  1.14 (0.88 – 1.48)  0.70 (0.30 – 1.61) | -  0.26  0.36  0.001  0.10  0.62  0.65  0.03  0.31  0.40 |
| **Time between questionnaire rollout and questionnaire completion (per day)** | 1.00 (1.00 – 1.00) | 0.04 | 1.00 (1.00 – 1.00) | 0.69 |
| **Occupational factors** | | | | |
| **Occupation**  Doctor or medical support  Nurse, nursing associate or Midwife  Allied health professional ^‡^  Dental  Admin, estates or other | Ref  1.40 (1.13 – 1.73)  1.12 (0.92 – 1.35)  0.87 (0.58 – 1.31)  1.37 (0.89 – 2.10) | -  0.002  0.26  0.24  0.16 | -  -  -  -  - | -  -  -  -  - |
| **Transport to work**  Alone or with members of household  With others outside household | Ref  1.05 (0.85 – 1.31) | -  0.65 | -  - | -  - |
| **Number of SARS-CoV-2 positive patients attended to per week (with physical contact)**  None  1 – 5  6 – 20  ≥ 21 | Ref  1.72 (1.44 – 2.07)  2.23 (1.80 – 2.75)  2.83 (2.14 – 3.73) | -  <0.001  <0.001  <0.001 | -  -  -  - | -  -  -  - |
| **Access to appropriate PPE**  Not applicable or all/most the time  Some of the time or less frequently | Ref  1.33 (1.16 – 1.52) | -  <0.001 | -  - | -  - |
| **Aerosol generating procedure exposure**  Less than weekly exposure  At least weekly exposure | Ref  0.91 (0.76 – 1.09) | -  0.30 | -  - | -  - |
| **Night shift pattern**  Never works nights  Works nights less than weekly  Works nights weekly or always | Ref  1.03 (0.86 – 1.25)  0.84 (0.67 – 1.06) | -  0.73  0.14 | -  -  - | -  -  - |
| **Work areas**  Ambulance  Community clinical setting /primary care  Non clinical community setting  Emergency Department  Intensive Care Unit  Hospital Inpatient  Hospital Outpatient  Hospital non-clinical area or laboratory  Psychiatric hospital  Maternity  Nursing or Care Home  University  Home | 2.02 (1.42 – 2.85)  0.92 (0.77 – 1.10)  0.91 (0.66 – 1.25)  1.09 (0.88 – 1.35)  0.73 (0.58 – 0.93)  1.58 (1.34 – 1.86)  0.95 (0.79 – 1.14)  0.88 (0.69 – 1.11)  1.31 (0.89 – 1.93)  0.72 (0.50 – 1.06)  1.46 (0.98 – 2.17)  0.97 (0.60 – 1.56)  0.84 (0.68 – 1.01) | <0.001  0.35  0.56  0.43  0.01  <0.001  0.58  0.29  0.17  0.09  0.06  0.89  0.08 | -  -  -  -  -  -  -  -  -  -  -  -  - | -  -  -  -  -  -  -  -  -  -  -  -  - |

Supplementary Table 6 shows the results of two multivariable logistic regression analyses analysing only those with no missing data in any variable of interest. These analyses examine the association of demographic and household factors with infection (in the larger cohort), and the other is additionally adjusted for occupational factors (in those working during lockdown).

*for each decade increase in age. † Wales, Scotland and Northern Ireland have been combined for this sensitivity analysis. This is because English IMD categories were imputed for those living in these nations. Therefore very few of those working in Scotland, Wales or Northern Ireland are included in this complete case analysis necessitating collapse of these levels of the work region variable into one level. ‡ Also includes pharmacists, healthcare scientists, ambulance workers and those in optical roles.

Analyses adjusted for all other variables in the table (with the exception of the exclusion of occupational predictors in the right hand columns – as indicated by the lack of results in the relevant sections).

All occupational factors (other than region of workplace) relate to work circumstances during the weeks following the first UK national lockdown on March 23^rd^ 2020. When asked about work areas participants could select multiple answers , therefore the work areas variables are ‘dummy’ variables comparing all those that did not select an area (reference) with all those that did. Region of workplace is included in the analysis of household and demographic factors as a proxy for the participants region of residence.

aOR – adjusted odds ratio, PPE – personal protective equipment, Ref – reference category for categorical variables, SARS-CoV-2 – severe acute respiratory syndrome coronavirus-2

**Supplementary Table 7. Univariable and multivariable analysis of factors associated with SARS-CoV-2 infection using a more granular ethnicity variable in those who worked during lockdown (n=10,772)**

| Variable | Univariable | | Multivariable | |
| --- | --- | --- | --- | --- |
|  | OR (95% CI) | p value | aOR (95% CI) | p value |
| **Age*** | 0.83 (0.80 – 0.86) | <0.001 | 0.92 (0.88 – 0.96) | <0.001 |
| **Sex**  Male  Female | Ref  0.94 (0.85 – 1.04) | -  0.25 | Ref  1.03 (0.91 – 1.16) | -  0.65 |
| **Ethnicity**  White - British  White - Irish  White - Other/Gypsy Irish Traveller  Asian - Indian  Asian - Pakistani  Asian - Bangladeshi  Asian - Chinese  Asian - Other  Black - African  Black - Caribbean  Black - Other  Mixed - White & Black Caribbean  Mixed - White & Black African  Mixed - White & Asian  Mixed - Other  Arab  Other | Ref  1.29 (0.94 – 1.79)  1.21 (1.02 – 1.43)  1.00 (0.86 – 1.17)  1.66 (1.29 – 2.13)  1.28 (0.73 – 2.23)  1.02 (0.74 – 1.39)  1.26 (0.98 – 1.60)  1.60 (1.26 – 2.04)  0.99 (0.63 – 1.55)  1.77 (0.71 – 4.38)  0.74 (0.41 - 1.32)  1.03 (0.57 – 1.87)  1.15 (0.80 – 1.64)  0.83 (0.54 – 1.28)  1.13 (0.73 – 1.76)  1.11 (0.72 – 1.71) | -  0.12  0.03  0.97  <0.001  0.39  0.92  0.07  <0.001  0.97  0.22  0.30  0.92  0.45  0.41  0.57  0.64 | Ref  1.35 (0.94 – 1.92)  1.19 (0.96 – 1.48)  0.88 (0.72 – 1.07)  1.21 (0.89 – 1.63)  0.97 (0.53 – 1.75)  0.90 (0.64 – 1.27)  0.92 (0.69 – 1.23)  1.20 (0.89 – 1.61)  0.79 (0.49 – 1.26)  1.46 (0.55 – 3.84)  0.65 (0.36 – 1.29)  0.98 (0.52 – 1.85)  1.01 (0.69 – 1.48)  0.81 (0.52 – 1.37)  0.84 (0.51 – 1.37)  0.96 (0.60 – 1.54) | -  0.10  0.11  0.19  0.22  0.91  0.56  0.58  0.24  0.32  0.45  0.16  0.96  0.96  0.37  0.48  0.86 |
| **Migration status**  Born in UK  Born abroad | Ref  1.16 (1.05 – 1.29) | -  0.003 | Ref  1.02 (0.87 – 1.18) | -  0.84 |
| **Religiosity**  Not important or not religious  Fairly important  Very important  Extremely important | Ref  1.05 (0.94 – 1.18)  1.12 (0.96 – 1.30)  1.33 (1.15 – 1.53) | -  0.40  0.16  <0.001 | Ref  1.07 (0.95 – 1.21)  1.03 (0.87 – 1.23)  1.25 (1.05 – 1.48) | -  0.14  0.71  0.01 |
| **Index of multiple deprivation**  1 (most deprived)  2  3  4  5 (least deprived) | 1.23 (1.02 – 1.47)  1.17 (1.00 – 1.36)  Ref  0.99 (0.85 – 1.14)  0.95 (0.82 – 1.09) | 0.03  0.04  -  0.86  0.45 | 1.02 (0.84 – 1.23)  1.08 (0.92 – 1.26)  Ref  1.02 (0.87 – 1.19)  1.05 (0.90 – 1.21) | 0.84  0.21  -  0.89  0.54 |
| **Household size** | 1.05 (1.02 – 1.08) | 0.004 | 1.01 (0.98 – 1.05) | 0.47 |
| **Cohabitation**  Does not live with other key workers  Lives with other key workers | Ref  1.29 (1.18 – 1.41) | -  <0.001 | Ref  1.17 (1.06 – 1.30) | -  0.002 |
| **Accommodation**  Does not have shared spaces  Has shared spaces | Ref  1.16 (1.04 – 1.30) | -  0.01 | Ref  0.92 (0.81 – 1.06) | -  0.25 |
| **Social mixing with others outside household**  None / remote only  Face to face with social distancing  With physical contact | Ref  0.82 (0.74 – 0.91)  1.02 (0.88 – 1.18) | -  <0.001  0.77 | Ref  0.90 (0.81 – 1.01)  0.99 (0.85 – 1.16) | -  0.07  0.93 |
| **Comorbidities**  Diabetes  Immunosuppression | 0.95 (0.75 – 1.21)  0.76 (0.57 – 1.00) | 0.68  0.06 | 1.09 (0.84 – 1.41)  0.99 (0.72 – 1.38) | 0.50  0.97 |
| **Shielding status**  Not advised to shield  Advised to shield | Ref  0.78 (0.60 – 1.00) | -  0.05 | Ref  0.86 (0.64 – 1.15) | -  0.32 |
| **Smoking status**  Ex or non-smoker  Current smoker | Ref  0.68 (0.54 – 0.85) | -  0.001 | Ref  0.56 (0.44 – 0.71) | -  <0.001 |
| **Time between questionnaire rollout and questionnaire completion (per day)** | 1.00 (1.00 – 1.00) | 0.73 | 1.00 (1.00 – 1.00) | 0.53 |
| **Occupation**  Doctor or medical support  Nurse, nursing associate or Midwife  Allied health professional†  Dental  Admin, estates or other | Ref  1.14 (1.00 – 1.30)  0.88 (0.79 – 0.99)  0.51 (0.38 – 0.68)  0.69 (0.55 – 0.86) | -  0.16  0.02  <0.001  0.001 | Ref  1.37 (1.17 – 1.61)  1.07 (0.93 – 1.24)  0.83 (0.61 – 1.14)  1.17 (0.90 – 1.51) | -  <0.001  0.35  0.25  0.23 |
| **Transport to work**  Alone or with members of household  With others outside household | Ref  1.39 (1.20 – 1.61) | -  <0.001 | Ref  1.10 (0.93 – 1.28) | -  0.26 |
| **Number of SARS-CoV-2 positive patients attended to per week (with physical contact)**  None  1 – 5  6 – 20  ≥ 21 | Ref  2.10 (1.87 – 2.35)  2.83 (2.49 – 3.20)  3.42 (2.89 – 4.04) | -  <0.001  <0.001  <0.001 | Ref  1.66 (1.45 – 1.90)  2.09 (1.79 – 2.45)  2.51 (2.05 – 3.08) | -  <0.001  <0.001  <0.001 |
| **Access to appropriate PPE**  Not applicable or all/most the time  Some of the time or less frequently | Ref  1.55 (1.41 – 1.70) | -  <0.001 | Ref  1.27 (1.15 – 1.40) | -  <0.001 |
| **Aerosol generating procedure exposure**  Less than weekly exposure  At least weekly exposure | Ref  1.48 (1.33 – 1.64) | -  <0.001 | Ref  0.90 (0.79 – 1.03) | -  0.12 |
| **Night shift pattern**  Never works nights  Works nights less than weekly  Works nights weekly or always | Ref  1.81 (1.61 – 2.02)  1.54 (1.35 – 1.76) | -  <0.001  <0.001 | Ref  1.10 (0.96 – 1.27)  0.85 (0.72 – 1.00) | -  0.17  0.06 |
| **Work areas**  Ambulance  Community clinical setting /primary care  Non clinical community setting  Emergency Department  Intensive Care Unit  Hospital Inpatient  Hospital Outpatient  Hospital non-clinical area or laboratory  Psychiatric hospital  Maternity  Nursing or Care Home  University  Home | 2.20 (1.79 – 2.71)  0.68 (0.61 – 0.76)  0.70 (0.56 – 0.87)  1.73 (1.50 – 2.00)  1.25 (1.08 – 1.46)  1.98 (1.80 – 2.19)  1.01 (0.90 – 1.14)  0.68 (0.58 – 0.80)  1.31 (1.02 – 1.68)  0.78 (0.59 – 1.03)  1.33 (1.00 – 1.76)  0.80 (0.57 – 1.12)  0.57 (0.50 – 0.66) | <0.001  <0.001  0.001  <0.001  0.03  <0.001  0.86  <0.001  0.04  0.08  0.05  0.20  <0.001 | 1.94 (1.51 – 2.50)  0.91 (0.80 – 1.04)  0.92 (0.73 – 1.17)  1.10 (0.94 – 1.30)  0.76 (0.63 – 0.91)  1.54 (1.36 – 1.73)  0.92 (0.80 – 1.05)  0.86 (0.72 – 1.02)  1.30 (0.99 – 1.69)  0.67 (0.50 – 0.89)  1.37 (1.02 – 1.85)  0.94 (0.66 – 1.34)  0.80 (0.69 – 0.93) | <0.001  0.19  0.51  0.23  0.003  <0.001  0.21  0.08  0.06  0.006  0.04  0.74  0.004 |
| **Work region**  West Midlands  London  South East England  South West England  East of England  East Midlands  North East England  North West England  Yorkshire and the Humber  Wales  Scotland  Northern Ireland | Ref  1.14 (0.95 – 1.39)  0.86 (0.70 – 1.05)  0.57 (0.45 – 0.73)  0.83 (0.66 – 1.04)  0.80 (0.65 – 0.99)  0.91 (0.69 – 1.18)  1.29 (1.06 – 1.58)  1.11 (0.89 – 1.39)  1.10 (0.83 – 1.46)  0.43 (0.32 – 0.57)  0.52 (0.31 – 0.87) | -  0.17  0.13  <0.001  0.10  0.04  0.47  0.01  0.35  0.49  <0.001  0.01 | Ref  1.13 (0.92 – 1.40)  0.85 (0.69 – 1.05)  0.59 (0.46 – 0.76)  0.81 (0.64 – 1.02)  0.86 (0.69 – 1.07)  0.87 (0.66 – 1.15)  1.22 (0.98 – 1.50)  1.15 (0.91 – 1.46)  1.13 (0.84 – 1.52)  0.42 (0.31 – 0.56)  0.49 (0.28 – 0.84) | -  0.24  0.14  <0.001  0.08  0.18  0.33  0.07  0.23  0.42  <0.001  0.009 |

Supplementary Table 5 shows the results of univariable and multivariable logistic regression analyses, examining the association of covariates with infection, in the cohort working during lockdown using the office or national statistics’ 18 ethnicity categories (with gypsy/Irish traveller collapsed into ‘White – Other’ due to low numbers of respondents in the former).

*for each decade increase in age. † Also includes pharmacists, healthcare scientists, ambulance workers and those in optical roles.

Analyses adjusted for all other variables in the table.

All occupational factors (other than region of workplace) relate to work circumstances during the weeks following the first UK national lockdown on March 23^rd^ 2020. When asked about work areas participants could select multiple answers , therefore the work areas variables are ‘dummy’ variables comparing all those that did not select an area (reference) with all those that did. Region of workplace is included in the analysis of household and demographic factors as a proxy for the participants region of residence.

aOR – adjusted odds ratio, PPE – personal protective equipment, Ref – reference category for categorical variables, SARS-CoV-2 – severe acute respiratory syndrome coronavirus-2
